# Evaluating an LLM-Assisted Workflow for Clinical Documentation: *A Pilot Randomized Controlled Trial on Time and Quality*

**DOI:** 10.1101/2025.10.06.25337211

**Authors:** Tomohiro Takayama, Keina Sado, Kenji Suda, Hiroshi Tamura, Naoko Ueda-Arakawa, Kenji Ishihara, Yuta Ueda, Kazuya Okamoto, Luciano Henrique de Oliveira Santos, Tetsuro Oshika, Tomohiro Kuroda, Akitaka Tsujikawa, Masahiro Miyake

## Abstract

**IMPORTANCE:** Large language models (LLMs) have been investigated for clinical documentation, with concerns about hallucinations and factual errors. Clinician review and revision of LLM-generated drafts are therefore considered essential, yet the impact of such workflow on both documentation time and quality remains unknown.

**OBJECTIVE:** To assess whether physician review and editing of LLM-generated drafts improves the time and quality of clinical documentation compared with clinician-only drafting in a randomized controlled trial setting.

**DESIGN:** Single-center, parallel-group, prospective, randomized, open-label, blinded-endpoint (PROBE) pilot trial conducted from February 18 to March 14, 2025.

**SETTING:** Kyoto University Hospital, Department of Ophthalmology.

**PARTICIPANTS:** Twenty-one ophthalmology physicians were randomized; 17 completed the study, and 4 withdrew before initiating intervention.

**INTERVENTIONS:** Participants were randomized to either the Clinician-in-the-loop group or the Clinician-only group. All participants created discharge summaries and referrals for six simulated patient records. In the Clinician-in-the-loop group, drafts were generated with an LLM assistant and then reviewed and edited by participants, whereas in the Clinician-only group, documents were drafted from scratch using matched templates. Unedited LLM drafts were additionally analyzed as the LLM-only group.

**MAIN OUTCOMES AND MEASURES:** Document creation time (primary) and expert-rated document quality across six domains plus overall quality (secondary).

**RESULTS:** Seventeen physicians submitted 48 discharge summaries and 48 discharge referrals in the Clinician-in-the-loop group, 54 of each document type in the Clinician-only group, and 48 of each in the LLM-only group. For summaries, Clinician-in-the-loop was associated with shorter creation time versus Clinician-only (β = −59.4 seconds; 95% CI, −118.1 to −0.8; P = .047). For referrals, clinician-in-the-loop required more time (β = 94.8 seconds; 95% CI, 40.4 to 149.3; P<.001). In most quality domains for both document types, the clinician-in-the-loop workflow outperformed clinician-only drafting. LLM-only drafts were fastest but had the lowest quality.

**CONCLUSIONS AND RELEVANCE:** A clinician-in-the-loop approach improved document quality and accelerated documentation. Active clinician review of LLM-generated drafts is essential for clinical documentation, and such workflows may help enhance working conditions and patient care.

**TRIAL REGISTRATION:** ClinicalTrials.gov Identifier NCT07187050

**Key Points:** *Question:* Does a workflow in which clinicians review and edit LLM-generated drafts (“clinician-in-the-loop”) improve the efficiency and quality of clinical documentation compared with clinician-only drafting?

*Findings:* In this single-center randomized controlled trial including 17 ophthalmology physicians, discharge summaries were significantly faster with the clinician-in-the-loop. Across both document types, clinician-in-the-loop drafts achieved higher quality scores than clinician-only documents.

*Meaning:* A clinician-in-the-loop workflow using LLMs can simultaneously enhance the efficiency and quality of clinical documentation; accordingly, active clinician review remains essential for improving working conditions and patient care overall.

## Introduction

Physicians devote a substantial proportion of their workday to clerical tasks, primarily electronic health record (EHR) documentation. A large U.S. time-motion study revealed that approximately 49% of physicians’ total working time is spent on EHR use and other desk work, with documentation alone accounting for roughly one-third of each patient encounter.^1^ Similarly, a survey of physicians at 80 Japanese hospitals found that EHR-related documentation consumed 13% of weekly working hours and represented one of the strongest predictors of overtime.^2^ This documentation workload not only extends working hours but also contributes to decreased job satisfaction and heightened risk of burnout among clinicians.^3–6^

Large language models (LLMs) have been proposed to mitigate this burden,^7^ which have received increasing attention because they can help simplify administrative tasks in healthcare.^8,9^ Recent investigations have applied models such as GPT 4 and Claude to the automated drafting of discharge summaries,^10–16^ discharge referrals,^16,17^ radiology reports,^18,19^ or other types of medical documents.^18,20–23^ However, comparative evaluations consistently report substantial limitations. Hallucinated findings,^11,13,21^ factual omissions,^21^ factual errors,^11–13,21,22^ and occasionally harmful recommendations,^13,19,22^ leading reviewers to conclude that LLM-generated documents are not sufficiently reliable for clinical practice.^13,19,22^

Because hallucinations and other factual inaccuracies are clinically unacceptable, LLM-generated drafts must be checked and corrected by clinicians.^15,20,24^ Accordingly, evaluation of the clinical utility of LLMs for document creation should consider the entire documentation process—a clinician-in-the-loop workflow that includes clinician review and editing after generation.^24^ However, most prior studies assess model output in isolation; to our knowledge, none has jointly evaluated time and quality within the workflow.

In the current study, to evaluate this clinician-in-the-loop workflow, we conducted a pilot randomized controlled trial using a novel LLM assistant, CocktailAI. This trial assessed documentation time and quality by comparing three approaches: documents drafted by our LLM assistant and then modified by clinicians, documents written entirely from scratch by clinicians, and documents generated by the LLM assistant with no human review.

## Methods

This study was reported following Consolidated Standards of Reporting Trials (CONSORT) reporting guideline for randomized clinical trials.^25^ The protocol for this study is provided in Supplement 1.

### Study Design and Setting

This single-center, parallel-group, prospective, randomized, open-label, blinded-endpoint (PROBE) trial was conducted at Kyoto University Hospital to evaluate the impact of the clinician-in-the-loop workflow. Participants were randomized to either an LLM-assisted documentation group (Clinician-in-the-loop group) or a manual, from-scratch documentation group (Clinician-only group). Randomization was performed using permuted block randomization with a block size of two and a 1:1 allocation ratio, stratified by participants’ position. The randomization list was generated in advance using an online tool (https://www.sealedenvelope.com/simple-randomiser/v1/lists), and participants were assigned sequentially according to this list. The study was conducted from February 18 to March 14, 2025. Participants were blinded to their group assignments prior to the initiation of the intervention; however, both participants and investigators were unblinded during the intervention phase. Outcome assessors evaluating the quality of the clinical documents were blinded to group assignments.

### Participants and Recruitment

Eligible participants included junior residents, senior residents, and attending ophthalmologists in the Department of Ophthalmology at Kyoto University Hospital. Physicians were eligible if they were actively working in the department during the study period and provide the confirmation that they did not routinely use CocktailAI for clinical documentation.

The study was conducted from February 18 to March 14, 2025. The investigators explained the study overview and eligibility criteria to the participants, and obtained written informed consent before any study procedures. Participants then completed a baseline questionnaire capturing demographic and professional characteristics, including age, gender, position, years of clinical experience, average daily time spent using EHRs, average number of days per week using EHRs, and the frequency of generative AI use.

### Simulated Medical Record

Six simulated patient records were developed in Japanese by Keina S. and Kenji S., representing diverse ophthalmology cases: cataract, ocular trauma, retinal detachment, corneal ulcer, optic neuritis, and glaucoma. Each record comprised comprehensive inpatient demographic data, pre-admission outpatient SOAP notes, and daily inpatient SOAP notes. The simulated records were formally approved by the Japanese Society of Artificial Intelligence in Ophthalmology as suitable for evaluation. English translations of all cases are provided in Supplement 2.

### Evaluation System

To precisely measure the time required for drafting and subsequent clinician modifications, we developed a purpose-built web application system that reproduces the clinical documentation workflow. In this system, CocktailAI, co-developed by the Department of Ophthalmology and Visual Sciences at Kyoto University Graduate School of Medicine, and Fitting Cloud Inc. (Kyoto, Japan), served as the LLM assistant. It assists in the creation of clinical documents by extracting relevant information from EHRs using LLMs and embedding the extracted content into predefined templates (Figure 1). In this study, the system employed the generative AI model Gemini-2.0-flash-lite for text generation.

**Figure 1.**
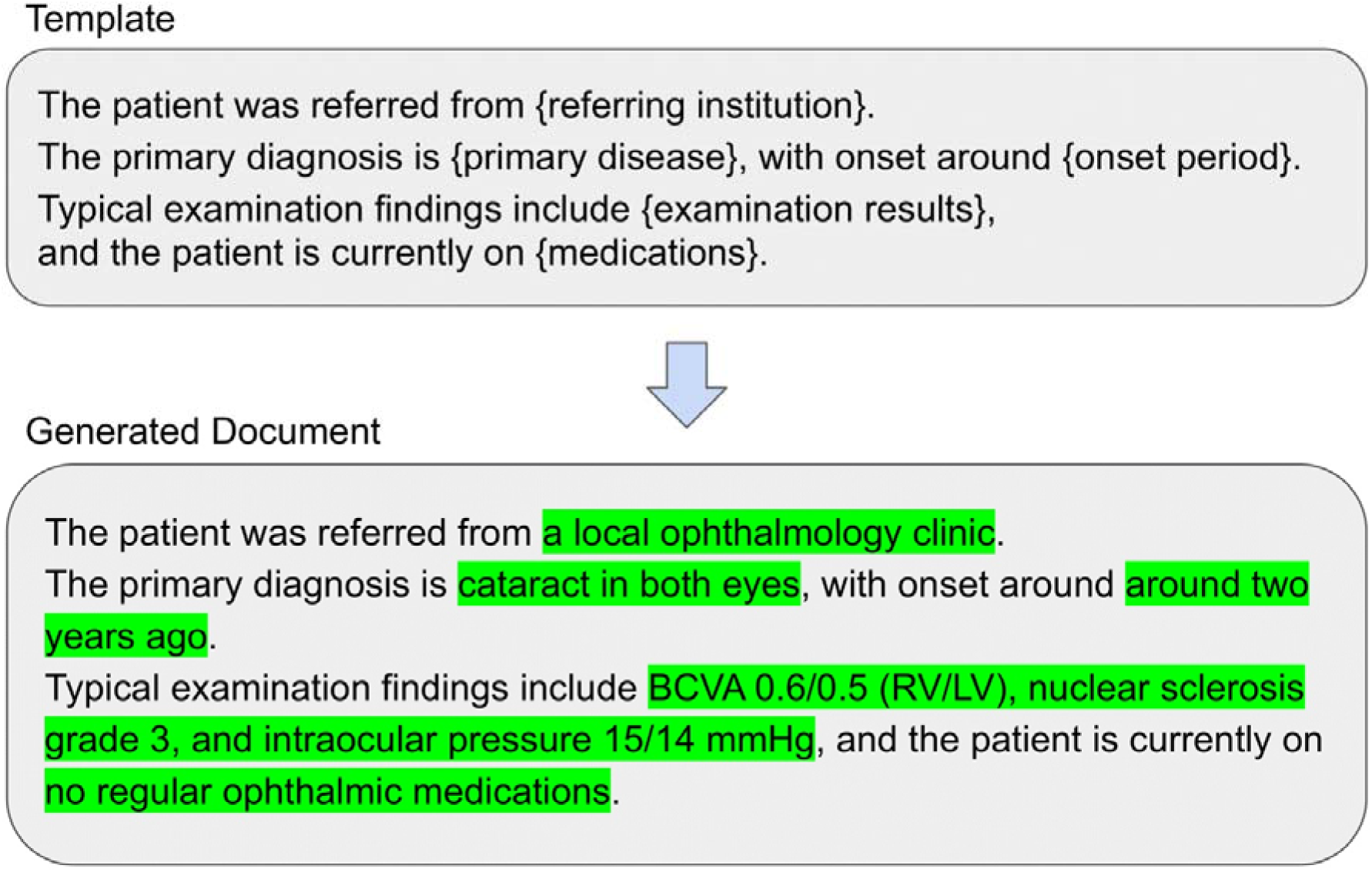
Example of Template-Based Document Generation. An LLM assistant was developed collaboratively by the Department of Ophthalmology and Visual Sciences at Kyoto University Graduate School of Medicine, and Fitting Cloud Inc. It extracts relevant information from EHRs and generates draft texts using predefined templates. Segments in {…} act as machine-readable directives, guiding the LLM assistant to retrieve the specified information directly from the EHR.

The templates used in this study were created by T.T., who was not involved in developing the simulated patient records. They were based on recent discharge summaries and discharge referrals from the Department of Ophthalmology. The complete content of each template is provided in Supplement 3.

### Study Arms

We tasked participants with creating two documents—discharge summary and discharge referral—for each of six simulated patient cases under two groups: Clinician-in-the-loop group and Clinician-only group. For each simulated case, participants created a discharge summary first and then a discharge referral; both the task order and the sequence of the six cases were fixed and identical for all participants.

In the Clinician-in-the-loop group, participants first generated a draft with the LLM assistant based on the predefined templates. Then, they reviewed the LLM-generated text and edited it.

Copy-and-paste from the patient record was permitted. No re-generation of LLM drafts was allowed.

In the Clinician-only group, participants worked from the same six patient records and completed the same two documents but had no access to the LLM assistant. They used templates with the same structure as those in the Clinician-in-the-loop group, although all LLM instruction prompts had been removed in advance to avoid the additional time needed to delete them for each use. Copy-and-paste from the record was also permitted.

Furthermore, an LLM-only group was included among the study groups. We retained the unedited drafts automatically produced by the LLM assistant before any human review or modification in the Clinician-in-the-loop group. These outputs represent LLM-generated documents without clinician review and were analyzed as a separate group.

The above information is presented in Figure 2.

**Figure 2.**
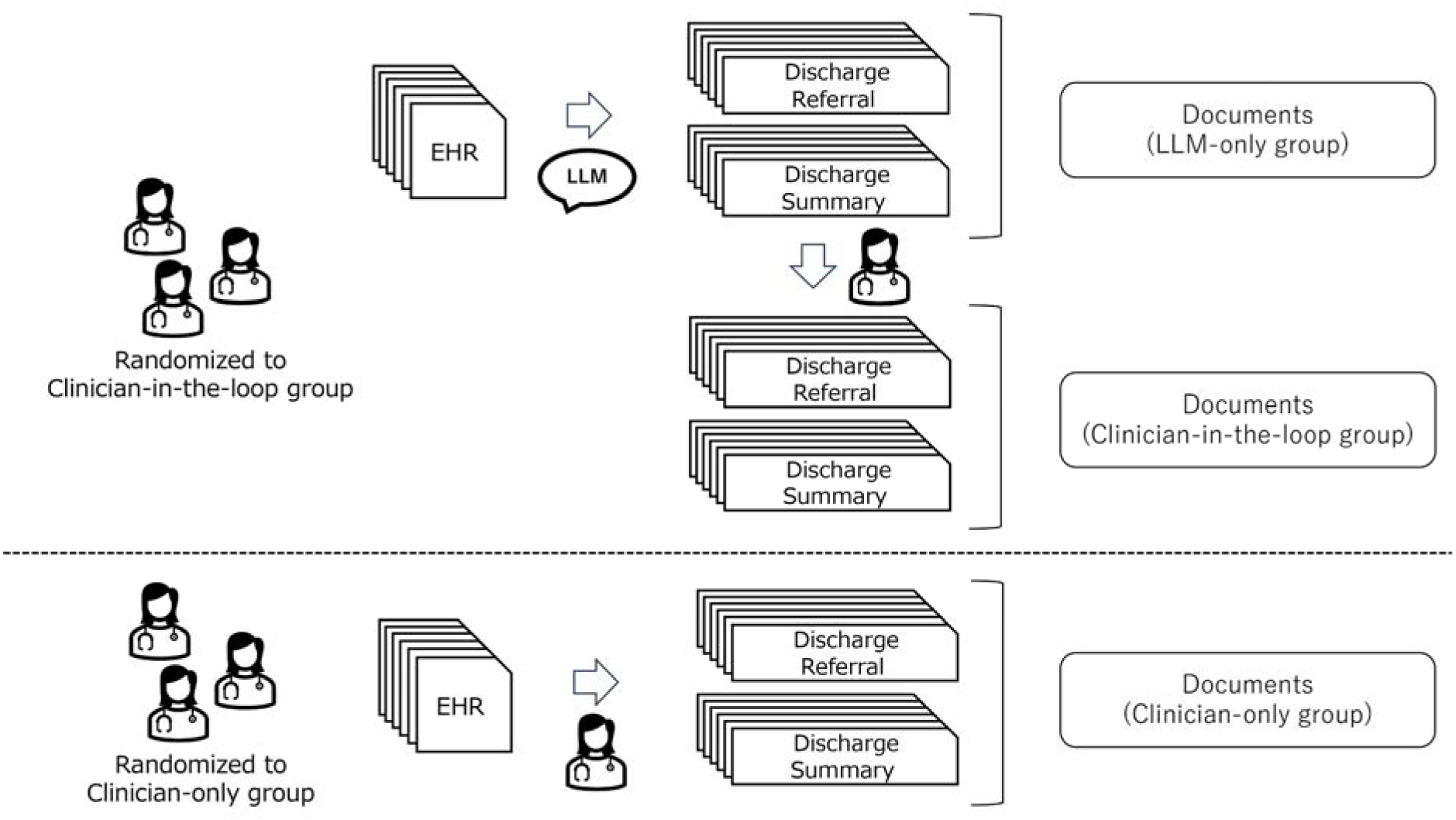
Study Arms and Clinical-Documentation Workflow across Six Simulated Cases. Participants were randomized to the Clinician-in-the-loop group or Clinician-only group. Clinician-in-the-loop group: participants used the LLM assistant to generate an initial draft, then edited the draft and submitted the final documents (discharge summary and discharge referral); in parallel, the unedited LLM drafts produced before any clinician review were saved and analyzed as an LLM-only group. Clinician-only group: For each of six simulated EHR cases, participants created and submitted the clinical documents directly from the EHR without the LLM assistant.

### Outcome Measures

The primary outcome was the average time required to complete each clinical document. Document creation time was automatically recorded within the study web application.

For the secondary outcome, document quality was independently evaluated by five ophthalmologists—Keina S. (Case 1), K.I. (Case 2), M.M. (Case 3), N.U.-A. (Case 4), and Kenji S. (Cases 5 and 6). Cases were matched to each evaluator’s clinical expertise. Evaluators were blinded to participant identity and study-arm. Document quality was assessed across six domains:

1. Medical Accuracy: The comprehensiveness of the information provided in the medical record
2. Language: Text style and vocabulary fit the situation and the intended target group that will read the text
3. Conciseness: The text contains only the strictly necessary information, and follows a logical or chronological order
4. Presence of Hallucinations: Any text that is factually incorrect or inconsistent, or unrelated to the medical record
5. Validity for Clinical Use: The text is deemed as usable in a true clinical scenario dealing with real patient information
6. Possibility of Harm: The severity and likelihood of potential harm based on people acting on the text

Domains 1–5 followed the framework of Sánchez-Rosenberg et al. (2024).^16^ Although Sánchez-Rosenberg et al. defined Domain 6 as “possibility of bias,” we instead applied the rubric of Singhal et al. (2023)^26^ and adapted Domain 6 to “possibility of harm,” since the former may suggest medical demographic bias, whereas our focus was on harmful medical misinformation. Detailed evaluation criteria —following Sánchez-Rosenberg et al. (2024) for Domains 1–5 and Singhal et al. (2023) for Domain 6—are provided in Supplement 4.

In addition to these domain-based evaluations, each document was subjectively rated on an overall quality scale from 0 to 10. Furthermore, evaluators were asked to guess which study group each document belonged to.

### Analysis

Although all participants were randomized, four participants withdrew before initiating any study procedures due to lack of time and provided no evaluable data; they had no exposure to the intervention and were unaware of their assignment. Consequently, analyses were conducted in the evaluable population, and an ITT analysis was not feasible because of missing data.

For the primary outcome, the mean and 95% confidence intervals were calculated separately for discharge summaries and discharge referrals. Separate linear mixed models (LMMs) were constructed for each document type (i.e., discharge summaries and discharge referrals) to analyze the primary outcome of document creation time. Specifically, group (the Clinician-only group vs. the Clinician-in-the-loop group) and clinical experience level (junior residents vs. all other participants) were included as fixed effects, and a random intercept was specified to account for the nested structure of cases within participants. As an exploratory analysis in this pilot study, an additional model including the group × clinical experience level interaction was fitted, and its estimates were reported alongside those of the primary models.

For the secondary outcome, descriptive statistics, including the mean for each evaluation domain, were calculated separately for each simulated patient case. In addition, for the overall quality scale, the mean and 95% confidence intervals were calculated for each study group, and for the task in which evaluators were asked to guess the group assignment of each document, corresponding correct identification rates were computed.

All analysis was implemented using SAS Studio version 3.81.

### Ethics

The Institutional Review Board of FINDEX Inc. reviewed this study and concluded that it was exempt from formal approval.

## Results

A total of 21 physicians were randomized (11 to Clinician-in-the-loop, 10 to Clinician-only). In the Clinician-in-the-loop group, 8 participants completed documentation tasks and 3 withdrew due to lack of time; in the Clinician-only group, 9 completed and 1 withdrew for the same reason. Because group assignment was revealed only after login to the study system, dropout occurred before participants knew their assignment. In total, 17 physicians completed the study and were included in the analysis. The Clinician-in-the-loop group submitted 48 discharge summaries and 48 referrals, and the Clinician-only group submitted 54 of each; 48 unedited LLM drafts were stored as the LLM-only group. The recruitment and allocation process is shown in Figure 3.

**Figure 3.**
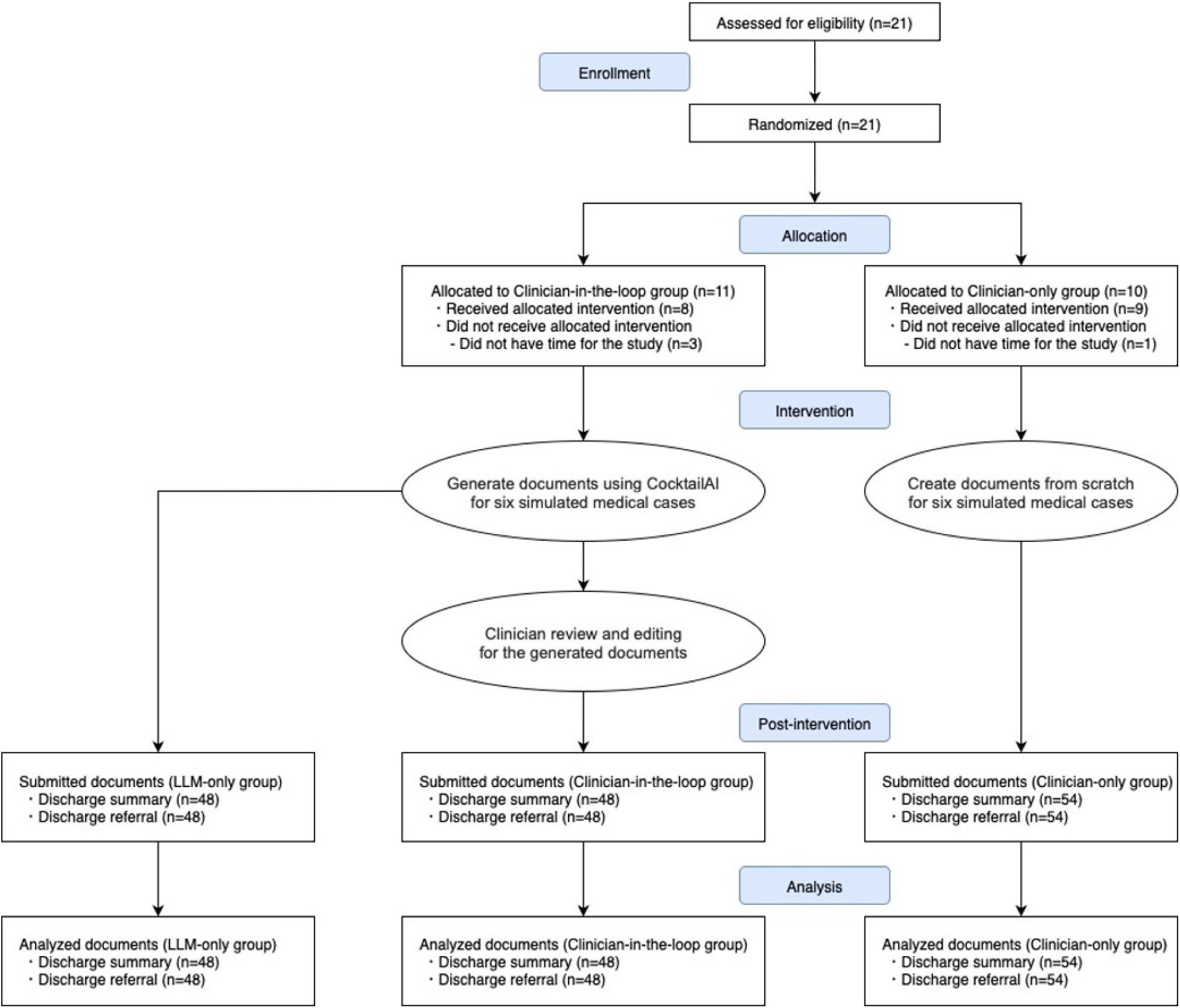
Participant Flowchart. A CONSORT-style flow diagram illustrating the enrollment, allocation, post-intervention, and analysis of participants.

Among the 17 participants, 35.3% were junior residents, 41.2% senior residents, and 23.5% attending ophthalmologists, with a mean clinical experience of 5.8 years. Other demographic information is provided in Table 1.

**Table 1.** Baseline Participant Demographics.

| **Participant Characteristic** | **Total (n=17)** | **Clinician-in-the-loop (n=8)** | **Clinician-only (n=9)** |
| --- | --- | --- | --- |
| Sex (Male), No. (%) | 12 (70.6) | 6 (75.0) | 6 (66.7) |
| Age, mean (SD), y | 30.5 (7.8) | 30.0 (5.1) | 31.0 (10.0) |
| Position, No. (%) |  |  |  |
| Junior Resident | 6 (35.3) | 3 (37.5) | 3 (33.3) |
| Senior Resident | 7 (41.2) | 3 (37.5) | 4 (44.4) |
| Attending Ophthalmologist | 4 (23.5) | 2 (25.0) | 2 (22.2) |
| Physician Experience, mean (SD), y | 5.8 (7.8) | 5.0 (5.5) | 6.4 (9.7) |
| EHR Use, mean (SD), hours/day | 3.7 (2.1) | 4.1 (2.3) | 3.3 (1.9) |
| EHR Use, mean (SD), days/week | 5.0 (1.8) | 4.9 (1.6) | 5.1 (2.1) |
| Generative AI Use, No. (%) |  |  |  |
| Never | 1 (5.9) | 0 (0.0) | 1 (11.1) |
| 0–1 days per week | 7 (41.2) | 2 (25.0) | 5 (55.6) |
| 1–3 days per week | 1 (5.9) | 1 (12.5) | 0 (0.0) |
| 3–5 days per week | 5 (29.4) | 3 (37.5) | 2 (22.2) |
| 5–7 days per week | 3 (17.6) | 2 (25.0) | 1 (11.1) |

### Primary Outcome

The average mean time required to complete discharge summaries and discharge referrals (measured in seconds) is illustrated in Figure 4. For discharge summaries, the Clinician-in-the-loop group demonstrated shorter mean times than the Clinician-only group in five of the six simulated cases. However, for discharge referrals, the Clinician-in-the-loop group required more time than the Clinician-only group in five of the six cases. A shorter mean time was observed only in Case 1. Detailed results are summarized in Table 2.

**Figure 4.**
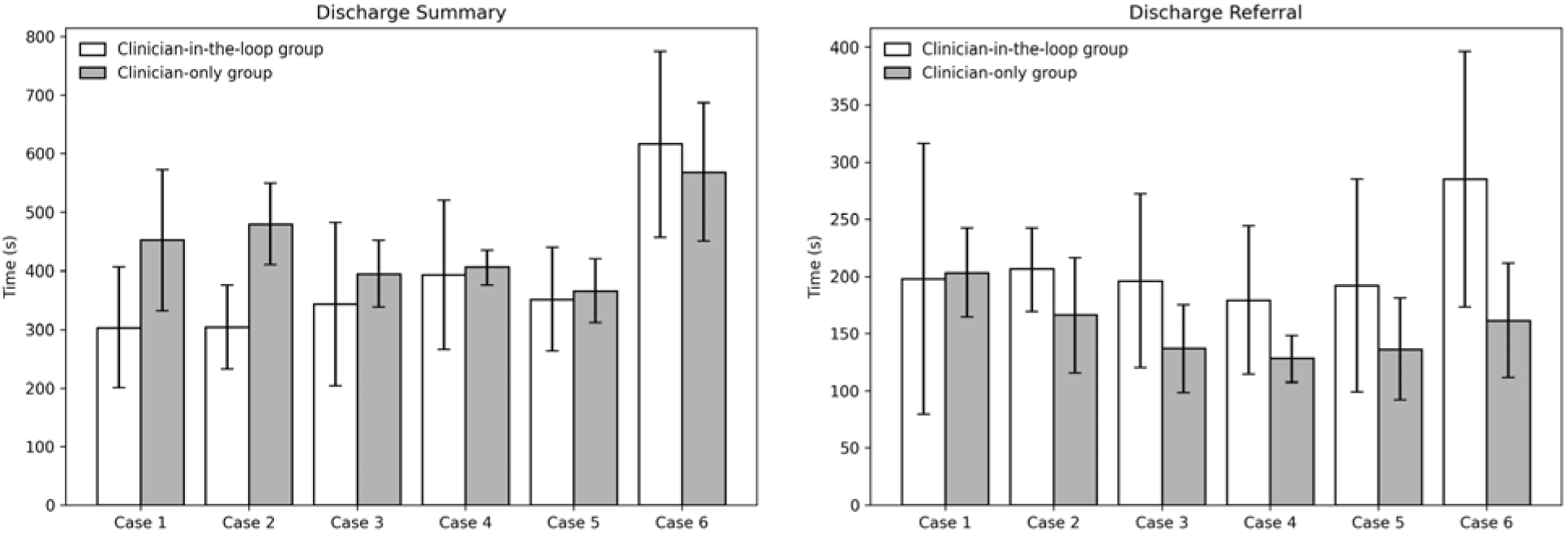
Time to Complete Clinical Documents Across Six Simulated Cases. Bar charts show the mean time (seconds) required to complete each document for six simulated patient cases, comparing the Clinician-in-the-loop group (orange) with Clinician-only group (blue). Left: Discharge summaries. Right: Discharge referrals. Error bars indicate 95% confidence intervals.

**Table 2.**
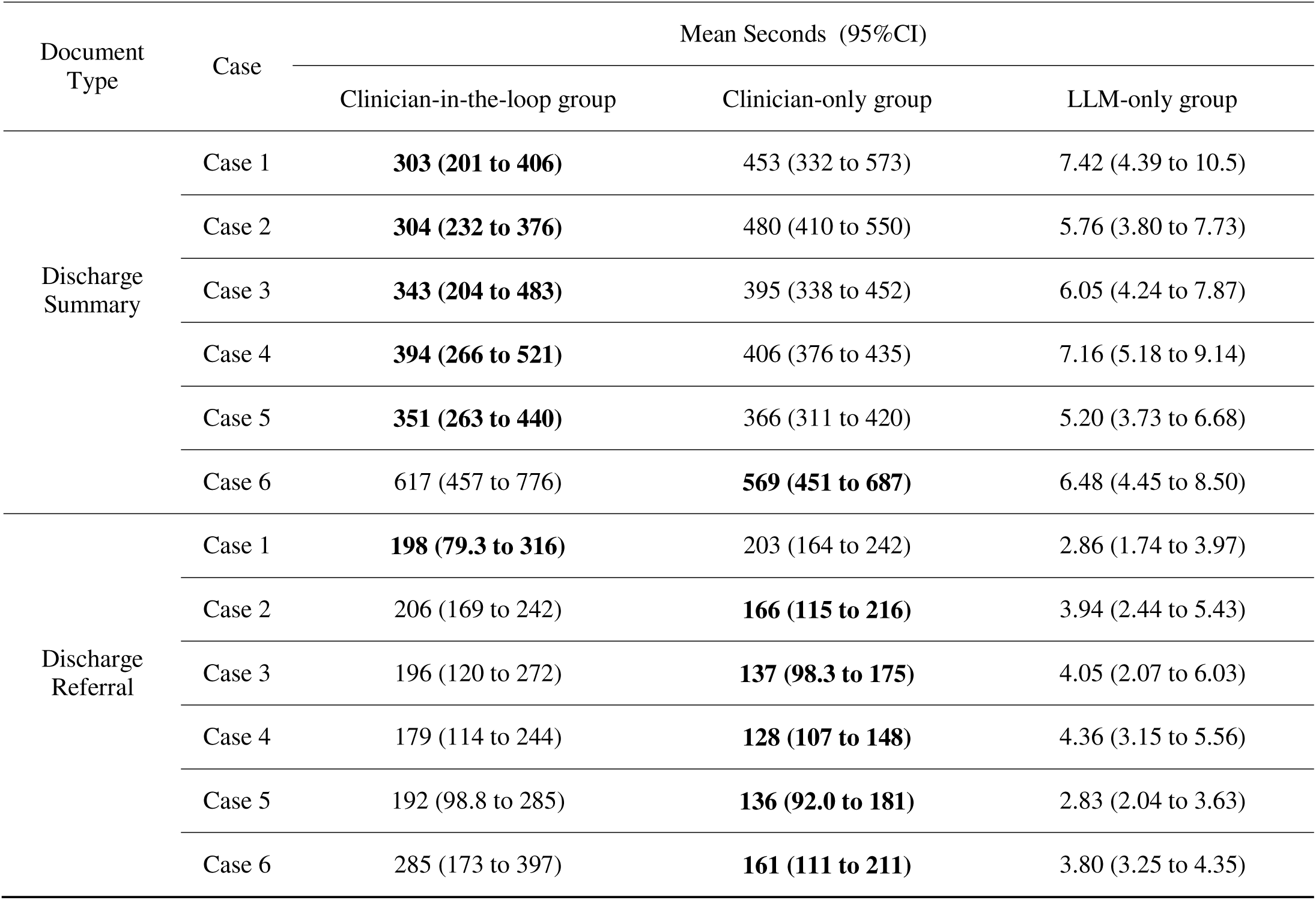
Mean Document Creation Time by Case and Group.

| Document Type | Case | Mean Seconds (95%CI) |  |  |
| --- | --- | --- | --- | --- |
|  |  | Clinician-in-the-loop group | Clinician-only group | LLM-only group |
| Discharge Summary | Case 1 | **303 (201 to 406)** | 453 (332 to 573) | 7.42 (4.39 to 10.5) |
|  | Case 2 | **304 (232 to 376)** | 480 (410 to 550) | 5.76 (3.80 to 7.73) |
|  | Case 3 | **343 (204 to 483)** | 395 (338 to 452) | 6.05 (4.24 to 7.87) |
|  | Case 4 | **394 (266 to 521)** | 406 (376 to 435) | 7.16 (5.18 to 9.14) |
|  | Case 5 | **351 (263 to 440)** | 366 (311 to 420) | 5.20 (3.73 to 6.68) |
|  | Case 6 | 617 (457 to 776) | **569 (451 to 687)** | 6.48 (4.45 to 8.50) |
| Discharge Referral | Case 1 | **198 (79.3 to 316)** | 203 (164 to 242) | 2.86 (1.74 to 3.97) |
|  | Case 2 | 206 (169 to 242) | **166 (115 to 216)** | 3.94 (2.44 to 5.43) |
|  | Case 3 | 196 (120 to 272) | **137 (98.3 to 175)** | 4.05 (2.07 to 6.03) |
|  | Case 4 | 179 (114 to 244) | **128 (107 to 148)** | 4.36 (3.15 to 5.56) |
|  | Case 5 | 192 (98.8 to 285) | **136 (92.0 to 181)** | 2.83 (2.04 to 3.63) |
|  | Case 6 | 285 (173 to 397) | **161 (111 to 211)** | 3.80 (3.25 to 4.35) |

Separate LMMs were constructed for discharge summaries and referrals to evaluate the effects of group assignment and clinical experience level on document creation time, with a random intercept for each participant-case combination. Full summaries of estimates are presented in Table 3.

**Table 3.** Linear Mixed Model Estimates for Document Creation Time.

| Document Type | Fixed Effect | Estimate ($\beta$) | 95% CI | Standard Error | P value |
| --- | --- | --- | --- | --- | --- |
| **LMM without Interaction Terms** |  |  |  |  |  |
| Discharge Summary | Intercept | 446.2 | 388.9 to 503.5 | 28.9 | <.001 |
|  | Group<sup>a</sup> | -59.4 | -118.1 to -0.7 | 29.6 | .047 |
|  | Clinical Experience Level<sup>b</sup> | -2.3 | -63.6 to 59.0 | 30.9 | .94 |
| Discharge Referral | Intercept | 170 | -124788 to 125128 | 62976 | >.99 |
|  | Group | 52.9 | -31474 to 31580 | 15889 | >.99 |
|  | Clinical Experience Level | -22.3 | -87528 to 87484 | 44101 | >.99 |
| **LMM with Interaction Terms** |  |  |  |  |  |
| Discharge Summary | Intercept | 441.0 | 371.0 to 511.0 | 35.3 | <.001 |
|  | Group | -49.1 | -148.1 to 50.0 | 49.9 | .33 |
|  | Clinical Experience Level | 5.5 | -80.2 to 91.2 | 43.2 | .90 |
| | Group × Clinical Experience Level (Interaction) | -16.0 | -139.3 to 107.2 | 62.1 | .80 |
| Discharge Referral | Intercept | 149.1 | 110.6 to 187.6 | 19.4 | <.001 |
|  | Group | 94.8 | 40.3 to 149.3 | 27.5 | <.001 |
|  | Clinical Experience Level | 9.2 | -38.0 to 56.3 | 23.8 | .70 |
| | Group × Clinical Experience Level (Interaction) | -64.9 | -132.8 to 2.9 | 34.2 | .060 |

a) Group: Clinician-only vs. Clinician-in-the-loop
b) Clinical Experience Level: junior residents vs. all other participants

In the model for discharge summaries, the Clinician-in-the-loop group assignment was associated with a significantly shorter document creation time compared to the Clinician-only group assignment (β = −59.4, 95% CI: −118.1 to −0.8, p = .047). No significant effect was observed for clinical experience level (β = −2.3, 95% CI: −63.6 to 59.0, p = .94).

In the additional model for discharge referrals, the Clinician-in-the-loop group assignment was significantly associated with longer document creation time (β = 94.8, 95% CI: 40.4 to 149.3, p<.001), while clinical experience level remained non-significant (p = .70). Although the interaction term between group and clinical experience level was not statistically significant (p = .060), the estimated value was −64.9 seconds (95% CI: −132.8 to 2.9).

### Secondary Outcome

The submitted clinical documents were evaluated across six predefined quality domains. The Clinician-in-the-loop group showed higher average scores than the Clinician-only group across nearly all domains in discharge summaries and discharge referrals. The LLM-only group consistently received the lowest scores across most domains for both discharge summaries and discharge referrals; however, in the discharge summaries the Possibility of Harm and Validity for Clinical Use domains, and in the discharge referrals the Possibility of Harm domain, achieved scores comparable to those of the Clinician-only group. (Figure 5).

**Figure 5.**
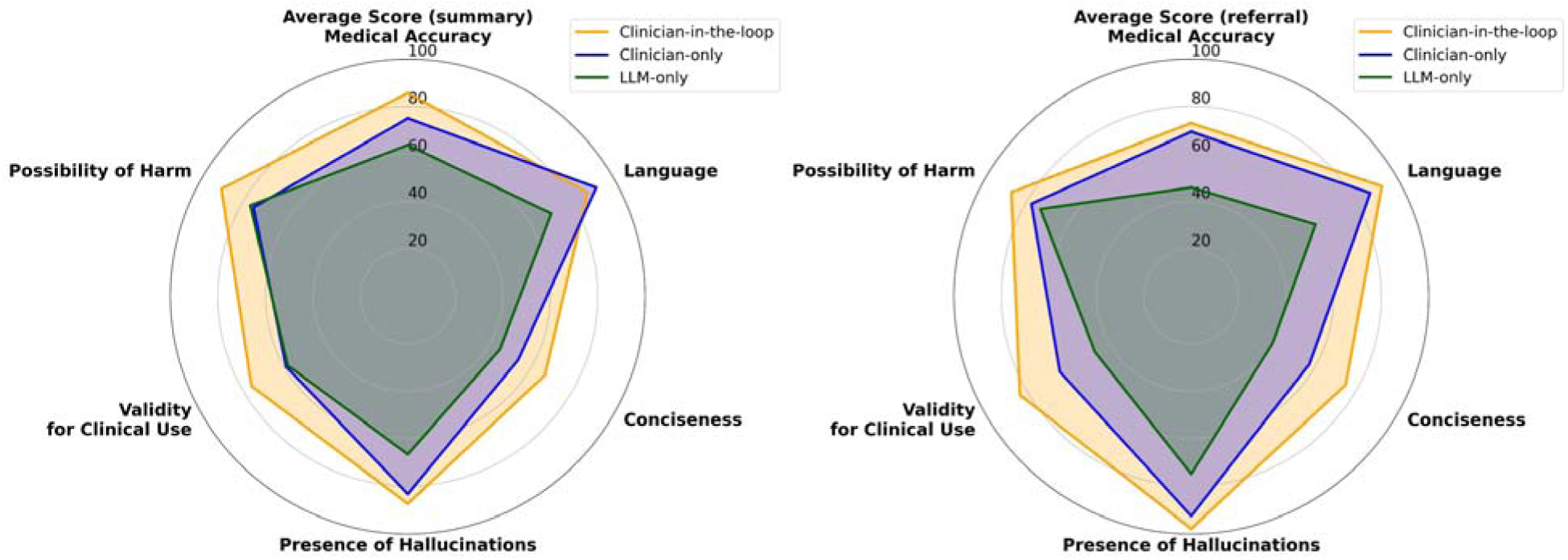
Average Evaluation Scores Across Document Types by Domain. Radar charts displaying the average evaluation scores for each quality domain across three groups (Clinician-in-the-loop, Clinician-only and LLM-only) for discharge summaries (left) and discharge referrals (right). Scores represent the mean values across all simulated cases, aggregated for six domains: Medical Accuracy, Language, Conciseness, Presence of Hallucinations, Validity for Clinical Use, and Possibility of Harm. Higher scores indicate better performance.

Each document was also rated on an overall quality scale from 0 to 10. The Clinician-in-the-loop group received the highest mean scores for both document types: 7.8 (95% CI: 7.4 to 8.2) for discharge summaries and 8.0 (95% CI: 7.6 to 8.4) for discharge referrals, respectively. Detailed results are summarized in Table 4.

**Table 4.** Overall Quality Ratings of Generated Documents by Group and Document Type.

| Document Type | Group | Mean Score | 95%CI |
| --- | --- | --- | --- |
| Discharge Summary | Clinician-in-the-loop | 7.8 | 7.4 to 8.2 |
|  | Clinician-only | 6.9 | 6.3 to 7.6 |
|  | LLM-only | 6.0 | 5.5 to 6.5 |
| Discharge Referral | Clinician-in-the-loop | 8.0 | 7.6 to 8.4 |
|  | Clinician-only | 6.8 | 6.2 to 7.4 |
|  | LLM-only | 5.3 | 4.9 to 5.8 |

For discharge summaries, the Clinician-in-the-loop group was correctly identified in 60.4% of cases, whereas the Clinician-only group was correctly identified in 75.9% of cases. For discharge referrals, the rates were 56.3% for the Clinician-in-the-loop group and 63.0% for the Clinician-only group (Table 5).

**Table 5.** Distribution of Predicted Group Assignments by Actual Study Group (%)

| Document Type | Actual Study Group | Predicted Group Assignment (%) |  |  |
| --- | --- | --- | --- | --- |
|  |  | Clinician-in-the-loop | Clinician-only | LLM-only |
| Discharge Summary | Clinician-in-the-loop | 60.4% (29/48) | 20.8% (10/48) | 18.8% (9/48) |
|  | Clinician-only | 16.7% (9/54) | 75.9% (41/54) | 7.4% (4/54) |
|  | LLM-only | 22.9% (11/48) | 2.1% (1/48) | 75.0% (36/48) |
| Discharge Referral | Clinician-in-the-loop | 56.3% (27/48) | 29.2% (14/48) | 14.6% (7/48) |
|  | Clinician-only | 18.5% (10/54) | 63.0% (34/54) | 18.5% (10/54) |
|  | LLM-only | 20.8% (10/48) | 0.0% (0/48) | 79.2% (38/48) |

## Discussion

This randomized controlled trial is the first to assess the real-world utility of LLMs for clinical documentation while explicitly evaluating both time and quality outcomes. We compared a clinician-in-the-loop workflow with manual drafting because LLM-only output remains difficult to implement safely in routine clinical practice. In the Clinician-in-the-loop group, physicians completed discharge summary more quickly, and clinician-edited LLM drafts consistently outperformed manually drafted documents across most quality domains and on overall expert-review scores. Exploratory analyses further suggested a potential interaction between clinician seniority and the effect of LLM use, offering a concrete implication for how clinician-in-the-loop approaches may deliver value in practice.

Several studies have compared AI-generated documents with those written by clinicians (Table 6). For relatively simple, short-form clinical texts—such as radiology reports and progress notes—LLM outputs have achieved performance comparable to human-written documents.^18^ However, as both the input prompts and target outputs lengthen (e.g., discharge referrals and discharge summaries), known issues such as hallucinations and other errors become more prominent.^11,12,15,16^ Consistent with these reports, our LLM-only group produced drafts far more quickly yet with inferior quality. In routine clinical care, such deficiencies are unacceptable; accordingly, any LLM-enabled documentation workflow should make clinician review and revision an essential step rather than an optional safeguard. This physician-led use of LLM is also preferred from the patient perspective, as shown by recent large-scale international survey data.^27^

**Table 6.**
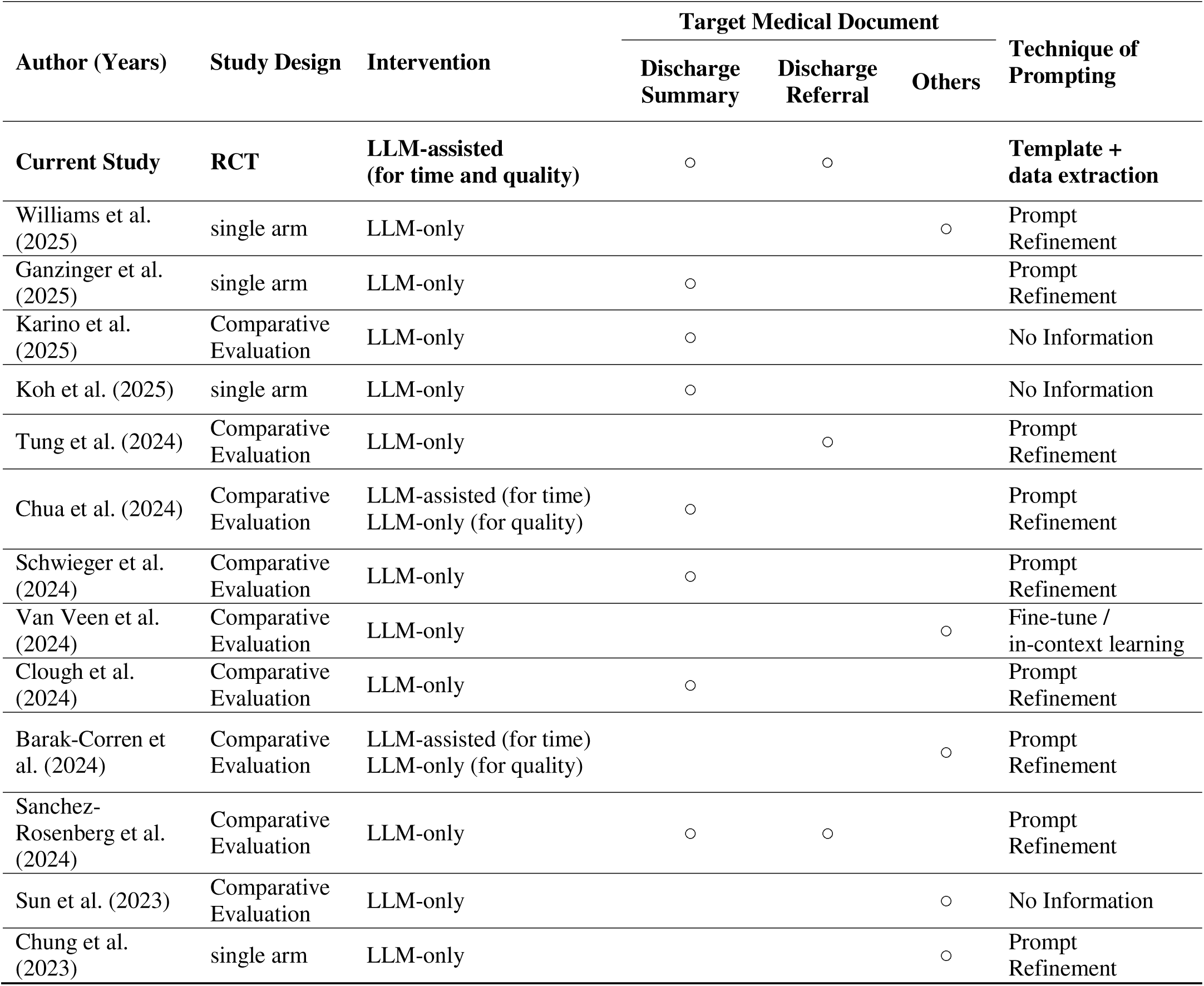
Overview of published studies evaluating LLM interventions for clinical document generation.

| Author (Years) | Study Design | Intervention | Target Medical Document |  |  | Technique of Prompting |
| --- | --- | --- | --- | --- | --- | --- |
|  |  |  | Discharge Summary | Discharge Referral | Others |  |
| **Current Study** | **RCT** | **LLM-assisted (for time and quality)** | ○ | ○ |  | **Template + data extraction** |
| Williams et al. (2025) | single arm | LLM-only |  |  | ○ | Prompt Refinement |
| Ganzinger et al. (2025) | single arm | LLM-only | ○ |  |  | Prompt Refinement |
| Karino et al. (2025) | Comparative Evaluation | LLM-only | ○ |  |  | No Information |
| Koh et al. (2025) | single arm | LLM-only | ○ |  |  | No Information |
| Tung et al. (2024) | Comparative Evaluation | LLM-only |  | ○ |  | Prompt Refinement |
| Chua et al. (2024) | Comparative Evaluation | LLM-assisted (for time)<br>LLM-only (for quality) | ○ |  |  | Prompt Refinement |
| Schwieger et al. (2024) | Comparative Evaluation | LLM-only | ○ |  |  | Prompt Refinement |
| Van Veen et al. (2024) | Comparative Evaluation | LLM-only |  |  | ○ | Fine-tune / in-context learning |
| Clough et al. (2024) | Comparative Evaluation | LLM-only | ○ |  |  | Prompt Refinement |
| Barak-Corren et al. (2024) | Comparative Evaluation | LLM-assisted (for time)<br>LLM-only (for quality) |  |  | ○ | Prompt Refinement |
| Sanchez-Rosenberg et al. (2024) | Comparative Evaluation | LLM-only | ○ | ○ |  | Prompt Refinement |
| Sun et al. (2023) | Comparative Evaluation | LLM-only |  |  | ○ | No Information |
| Chung et al. (2023) | single arm | LLM-only |  |  | ○ | Prompt Refinement |

Two studies have evaluated documentation time in clinician-in-the-loop workflows, one focused on discharge summaries and one on supervisory notes.^15,23^ Both reported reduced completion time which aligns with our findings. However, neither study assessed document quality, leaving unresolved whether efficiency gains were achieved at the expense of accuracy or completeness. This is the first study to jointly evaluate documentation time and quality, and the first to assess the real-world clinical use of LLM-assisted documentation in a randomized controlled trial. Across multiple quality domains, the clinician-in-the-loop approach outperformed manual drafting, indicating that this method is not merely “quick and dirty” but instead improves quality while reducing documentation burden. These results suggest that clinician-in-the-loop deployment of LLMs can simultaneously raise document quality and relieve workload, a combination that may translate into better patient care overall.

In this study, we implemented an extraction-based system and deployed it as an LLM assistant, reflecting the evidence that her data extraction can be highly accurate^28–30^. The observed difference in completion times between discharge summaries and discharge referrals likely reflects differences in document structure. For highly standardized documents such as discharge summaries, direct extraction into fixed templates may be sufficient. By contrast, discharge referrals involve greater narrative variability, and documents created from rigid templates often require additional modification to fit individual physician preferences. When creating documents with such narrative diversity, providing support for flexible, clinician-authored template creation alongside structured data extraction may be more appropriate and is likely to facilitate broader clinical adoption and accelerate real-world implementation.

We also examined heterogeneity by clinician seniority during LLM use. The interaction between clinician experience and the effects of AI assistance has been investigated in the recognition of radiological and pathological images,^31–36^ whereas, to our knowledge, no such evidence has been available in the context of LLMs. A sizable estimated seniority-group interaction in our study provides the first evidence in clinical LLM applications that greater clinical expertise may amplify the benefits of assistance. This finding suggests that LLM-specific training alone is unlikely to be sufficient; deeper clinical expertise may be required to maximize the benefits of LLMs in documentation workflows. Beyond documentation, AI in medicine now extends to areas such as clinical reasoning,^37^ image recognition,^38,39^ and basic research,^40^ and it is important to investigate whether similar patterns are observed in these domains.

This study has several limitations. First, our results reflect the capabilities of current language models. Future models with enhanced capabilities may produce different outcomes; nonetheless, our trial establishes a baseline for current model performance herEHR-focused applications. Second, we tested the system only in ophthalmology. Further research across other medical specialties is needed, including hospital-wide assessments of time savings, documentation quality, and downstream clinical outcomes. Third, templates were authored by an investigator rather than participants, so they may not fully reflect individual writing styles, as discussed earlier. Although we derived templates from recent discharge summaries and referrals in our Ophthalmology department to minimize stylistic mismatch, inter-physician variability likely persisted and may have influenced results. Future studies should evaluate the LLM system using clinician-authored, personalized templates prior to testing.

In conclusion, a clinician-in-the-loop workflow both accelerated documentation and improved overall document quality. Accordingly, successful implementation of AI tools in clinical practice requires active clinician supervision. When hospitals combine accurate AI-generated drafts with thorough human review, they can achieve higher quality documentation, shorter writing times, and, as a consequence, improved working conditions and patient care overall.

## Abbreviations

LLM: Large Language Model
EHR: Electronic Health Record
CI: Confidence Intervals
LMM: Linear Mixed Model

## Author Contribution

T.T. and Keina S. had full access to all the data in the study and took responsibility for the integrity of the data and the accuracy of the data analysis.

Conceptualization and Study design: T.T., Keina S., Kenji S., H.T., M.M.

Development of a purpose-built web application system: T.T., K.O., L.S.

Development of the simulated medical record: Keina S., Kenji S., T.O., M.M.

Data Collection: T.T.

Assessment of document quality: Keina S., Kenji S., N.U.-A., K.I., M.M.

Statistical analysis: T.T., Keina S., Y.U.

Interpretation of Results: T.T., Keina S., Kenji S., H.T., M.M.

Drafting of the manuscript: T.T.

Critical revision of the manuscript for important intellectual content: all authors.

Supervision: T.K., A.T., M.M.

## Disclaimer

Responsibility for the study’s analyses, findings, and opinions rests entirely with the authors. The content does not reflect the funder’s official policy or position, and no endorsement by the funder is implied.

## Declaration of generative AI use in the writing process

During the preparation of this work, the authors used ChatGPT (GPT-4o and GPT-5 by OpenAI) for translation, readability improvement, and proofreading of the English text. After using these services, the authors reviewed and edited the content as needed and took full responsibility for the content of the publication.

## Data Availability

The simulated patient records created for this study are publicly available in Supplement 2. Templates for the discharge summary and the discharge referral are provided in Supplement 3. Detailed evaluation criteria—following Sánchez-Rosenberg et al. (2024) for Domains 1–5 and Singhal et al. (2023) for Domain 6—are provided in Supplement 4.

