## Supplement 1 for "Evaluating an LLM-Assisted Workflow for Clinical Documentation: *A Pilot Randomized Controlled Trial on Time and Quality*"

**Protocol**

**Study Identification**

**Brief Title:**

LLM-Assisted vs Manual Writing for Clinical Documentation: Effects on Time and Quality

**Study Type:** Interventional

**Official Title:**

Evaluating an LLM-Assisted Workflow for Clinical Documentation: A Pilot Randomized Controlled Trial on Time and Quality

**Study Status**

**Record Verification Date:** August 2025

**Overall Recruitment Status:** Completed

**Actual Study Start Date:** February 18 2025

**Actual Primary Completion Date:** March 14 2025

**Actual Study Completion Date:** July 16 2025

**Sponsors and Collaborators**

**Responsible Party, by Official Title:** Principal Investigator

**Investigator Name:** Tomohiro Takayama

**Investigator Official Title:** Principal Investigator

**Investigator Affiliation:** Kyoto University, Graduate School of Medicine

**Name of the Sponsor:** Kyoto University, Graduate School of Medicine

**Collaborators:** Fitting Cloud Inc.

**Oversight**

**U.S. FDA-regulated Drug:** No

**U.S. FDA-regulated Device:** No

**U.S. FDA IND/IDE:** No

**Human Subjects Protection Review Board Status:** Exempt

**Data Monitoring Committee:** No

**Study Description**

**Brief Summary:**

The goal of this clinical trial is to learn whether an LLM-assisted writing workflow can reduce the time to complete hospital discharge summaries and discharge referrals and maintain or improve document quality compared with writing from scratch by clinicians in the Department of Ophthalmology at Kyoto University Hospital. The study used six simulated patient records (no real patient data).

The main questions it aims to answer are:

- Does the LLM-assisted writing workflow reduce the time needed to complete each document compared with manual writing?
- Does the LLM-assisted writing workflow maintain or improve document quality compared with manual writing, as rated by blinded experts?

Researchers will compare LLM-assisted versus manual writing to see if the LLM-assisted approach is faster and has equal or better quality. LLM-only drafts (unedited first drafts) will be evaluated as a separate third group to understand the baseline quality of LLM output without clinician edits.

Participants will create two documents—a discharge summary and a discharge referral—for each of six simulated cases. Those assigned to CocktailAI & Modification group will use an LLM assistant (called CocktailAI) to generate a first draft for each document and then review and edit it to finalize; those assigned to the control group will write each document from scratch without LLM assistance.

**Study Design**

**Primary Purpose:** Health Services Research

**Study Phase:** N/A

**Interventional Study Model:** Parallel Assignment

**Model Description:** This is a prospective, randomized, open-label, blinded-endpoint (PROBE) trial.

**Number of Arms:** 3

**Masking:** Single (Outcomes Assessor)

**Allocation:** Randomized

**Actual Enrollment:** 21

**Arms and Interventions**

| **Arms** | **Interventions** |
| --- | --- |
| **Experimental : CocktailAI & Modification arm**  Participants log into the study web application using a unique ID and their name. For each simulated patient case, they press “Start” to unlock the case record and initiate timing. After viewing the patient information, clinicians use CocktailAI to generate a first draft of the discharge summary. They then review and edit the draft and submit the final document by pressing “Submit.” For the discharge referral of the same case, clinicians follow the same procedure. Copy-and-paste from the case record is permitted. This sequence is repeated for six simulated cases. | **Other: Template-Based LLM Assistant**  This study uses CocktailAI, a template-based LLM assistant co-developed by the Department of Ophthalmology and Visual Sciences, Kyoto University Graduate School of Medicine, and Fitting Cloud Inc. (Kyoto, Japan). It is designed to extract relevant information from EHRs using LLMs and embed the extracted content into predefined templates. In this trial, the inputs are six simulated patient records (no real patient data). Text generation uses Gemini-2.0-flash-lite. Templates for discharge summaries and discharge referrals are pre-defined by a team member. |
| **Active Comparator : Control arm**  Participants log into the study web application using a unique ID and their name. For each simulated patient case, they press “Start” to unlock the case record and initiate timing. After viewing the patient information, clinicians write the discharge summary from scratch and submit the document by pressing “Submit.” They then write the discharge referral for the same case from scratch and submit it. Copy-and-paste from the case record is permitted. This sequence is repeated for six simulated cases. | **Other: Manual Writing**  The same document templates are provided; however, all LLM instruction prompts are removed in advance. Clinicians manually write the documents, following the template structure, for each of the six simulated cases. |
| **Active Comparator : CocktailAI arm**  The LLM assistant generates drafts of discharge summaries and discharge referrals directly from the simulated patient records, with no clinician review or edits. These drafts are the unedited first drafts produced in the CocktailAI & Modification arm (captured before any clinician edits) and are included as a separate group. | **Other: Template-Based LLM Assistant**  This study uses CocktailAI, a template-based LLM assistant co-developed by the Department of Ophthalmology and Visual Sciences, Kyoto University Graduate School of Medicine, and Fitting Cloud Inc. (Kyoto, Japan). It is designed to extract relevant information from EHRs using LLMs and embed the extracted content into predefined templates. In this trial, the inputs are six simulated patient records (no real patient data). Text generation uses Gemini-2.0-flash-lite. Templates for discharge summaries and discharge referrals are pre-defined by a team member. |

**Outcome Measures**

**Primary Outcome Measures**: Average time spent creating each document

[Time Frame: On one study day within 2 weeks after enrollment]

In the CocktailAI & Modification group and the Control group, the time spent creating documents is measured in seconds. In the CocktailAI group, the time required for document generation is measured in seconds.

**Secondary Outcome Measures**: Document quality assessment

[Time Frame: On one study day within 2 weeks after enrollment]

Blinded to group allocation, ophthalmology experts evaluate the documents using pre-specified criteria defined before study initiation. These criteria are developed based on six domains: Medical Accuracy, Language, Conciseness, Presence of Hallucinations, Validity for Clinical Use, and Possibility of Harm. Most domains are assessed on a three- point scale, whereas Presence of Hallucinations is evaluated dichotomously (present or absent). In addition to these domain-specific ratings, experts provide a subjective overall score on a 10-point scale (with higher scores indicating better quality) and are asked to guess which study group the document belongs to.

**Eligibility**

**Accepts Healthy Volunteers:** No

**Sex:** All

**Age Limits:** No limits

**Eligibility Criteria**

Inclusion Criteria:

- Ophthalmologists at Kyoto University Hospital
- Junior residents, senior residents, graduate students, board-certified ophthalmologists
- Physicians who confirm that they do not routinely use CocktailAI for clinical documentation and provide informed consent after receiving an explanation of the study

**Contacts and Locations**

**Central Contact Person:**

Tomohiro Takayama

**Central Contact Backup:**

Masahiro Miyake

**Locations:**

Department of Ophthalmology and Visual Sciences Kyoto University Graduate School of Medicine 54 Shogoin, Kawahara, Sakyo

Kyoto, Japan 606-8507

**Principal Investigator:**

Tomohiro Takayama, Keina Sado, Kenji Suda, Hiroshi Tamura, Masahiro Miyake
