## Supplement 2 for "Evaluating an LLM-Assisted Workflow for Clinical Documentation: *A Pilot Randomized Controlled Trial on Time and Quality*"

**The Six Simulated Patient Records**

**Case 1: Elderly Female Presenting for Cataract Surgery**

**Patient Name**: Kyoto Miyako
**Gender**: Female
**Age**: 77 years
**Date of First Visit**: January 4, 2024

**Diagnosis**

1. Bilateral nuclear cataracts
2. Bilateral hyperopic astigmatism

**History of Present Illness**

The patient, a 77-year-old female, has experienced progressive visual deterioration and photophobia in both eyes over the past 10 years. She consulted a local ophthalmologist, who diagnosed bilateral nuclear cataracts and hyperopic astigmatism and referred her to our hospital for further evaluation and treatment.

**Past Medical History**

- Essential hypertension

**Current Medication**

Amlodipine Besilate　5 mg (1 tablet daily in the morning)

**Allergies**

- None

**Social History**

The patient is fully independent in her activities of daily living (ADLs). She is a homemaker living with her husband, who has mild dementia and is under her care.

**Subjective Information**

"The previous doctor recommended surgery."

**Objective Information**

- **Visual Acuity**:
  - Right eye (RV): 0.4 (improves to 0.7 with S +2.00D, C -0.75D Ax 100)
  - Left eye (LV): 0.5 (improves to 0.6 with S +1.75D, C -0.75D Ax 115)
- **Intraocular Pressure**: 13 mmHg in both eyes (measured by NCT)
- **Axial Length**:
  - Right eye: 22.57 mm
  - Left eye: 22.54 mm
- **Anterior Chamber Depth**:
  - Right eye: 3.17 mm
  - Left eye: 3.09 mm
- **Corneal Endothelial Cell Density**:
  - Right eye: 3038 cells/mm²
  - Left eye: 2831 cells/mm²
- **Pupillary light reflex:** Present bilaterally (+/+)
- **RAPD**: Absent (-)
- **Ocular motility**: Normal
- **Slit Lamp Examination**:
  - **Conjunctiva**: No pathological findings
  - **Cornea**: Clear
  - **Anterior chamber**: Deep and quiet (cell -)
  - **Iris**: Round and smooth
  - **Lens**: Grade 2 nuclear sclerosis (NS2), anterior capsule opacification (ACO+), posterior subcapsular opacification absent (PSCO-)
  - **Fundus**: Well-visible and attached
- **Fundus Photography**: No abnormalities detected
- **OCT Findings**: No abnormalities detected

**Assessment**

1. Bilateral nuclear cataracts
2. Bilateral hyperopic astigmatism

**Plan**

Hospital admission for bilateral cataract surgery is planned.

- **Planned Admission Date**: February 1, 2024
- **Planned Surgeries**:
  - Right eye: February 1, 2024
  - Left eye: February 2, 2024
- **Discharge Date**: February 3, 2024

**Procedure**: Phacoemulsification with intraocular lens (IOL) insertion (Alcon CNA0T0 IOLs), targeting emmetropia.

- IOL power: +22.5D for both eyes

**Postoperative Follow-up**: The patient will return to the hospital for follow-up on postoperative days 3 and 7. If no complications are observed, care will be transferred back to her original ophthalmologist.

**Progress Notes**

**Hospital Day #1: February 1, 2024**

**Preoperative Examination**:

**Subjective**: "I feel fine as usual."

**Objective**:

- **Slit-lamp Findings**:
  - **Conjunctiva**: No pathological findings
  - **Cornea**: Clear
  - **Anterior Chamber**: Deep and quiet (cell -)
  - **Iris**: Round and smooth
  - **Lens**: NS2 ACO+ PSCO+-
  - **Fundus**: Well-visible and attached

**Assessment**: No abnormalities detected preoperatively.
**Plan**: Perform phacoemulsification and IOL insertion for the right eye today.

**Hospital Day #2: February 2, 2024**

**Postoperative Day 1 (Right Eye)**:

**Subjective**: "I can see better with my right eye. There’s no pain."

**Objective**:

- **Intraocular Pressure**: RT 12 mmHg / LT 10 mmHg
- **Right Eye**:
  - **Conjunctiva**: Hyperemia (+), Papilla (-) Follicle (-)
  - **Cornea**: Clear, no wound leakage
  - **Anterior Chamber**: Deep, cell (+)
  - **Iris**: Round and smooth
  - **IOL**: In the capsular bag
  - **Fundus**: Well-visible and attached
- **Left Eye**:
  - **Conjunctiva**: No pathological findings
  - **Cornea**: Clear
  - **Anterior Chamber**: Deep, cell (-)
  - **Iris**: Round and smooth
  - **Lens**: NS2 ACO+ PSCO+-
  - **Fundus**: Well-visible and attached

**Assessment**:

- Good postoperative progress in the right eye
- No abnormalities in the left eye

**Plan**: Perform phacoemulsification and IOL insertion for the left eye today.

**Postoperative Medications for the Right Eye**:

1. Levofloxacin Hydrate (4x/day)
2. Fluorometholone (4x/day)
3. Bromfenac Sodium Hydrate(2x/day)

**Postoperative Informed Consent (IC):**

- During the left-eye PEA + IOL procedure, zonular fragility and lens instability were observed. A posterior capsular rupture occurred during ultrasonic emulsification of the lens nucleus. The remaining nuclear and cortical fragments were carefully aspirated without evidence of lens material dropping into the vitreous cavity.
- An anterior vitrectomy was performed to address the situation, ensuring no vitreous prolapse was present. An NS70 +21.5 intraocular lens (IOL) was securely placed *on the bag*.
- The absence of vitreous incarceration was confirmed, and the surgery was concluded without further complications.
- The findings and surgical course were explained to the patient, who demonstrated good understanding and acknowledged the explanation with a clear, "I understand."

**Hospital Day #3: February 3, 2024**

**Postoperative Day 2 (Right Eye), Day 1 (Left Eye)**:

**Subjective**: "My right eye is much clearer, but my left eye is still blurry."

**Objective**:

- **Visual Acuity**:
  - Right eye: 0.7 (improves to 1.5x with S -0.25D, C -0.50D Ax 145)
  - Left eye: 0.3 (improves to 0.5x with S -0.50D, C -0.75D Ax 125)
- **Intraocular Pressure**: RT 10 mmHg / LT 15 mmHg (measured by applanation)
- **Right Eye**:
  - **Conjunctiva**: Hyperemia (+), Papilla (-) Follicle (-)
  - **Cornea**: Clear, no wound leakage
  - **Anterior Chamber**: Deep, cell (+)
  - **Iris**: Round and smooth
  - **IOL**: In the capsular bag
  - **Fundus**: Well-visible and attached
- **Left Eye**:
  - **Conjunctiva**: Hyperemia (+), Papilla (-) Follicle (-)
  - **Cornea**: Clear, no wound leakage, Descemet’s folds (DF++)
  - **Anterior Chamber**: Deep, cell (++), small fibrin deposits
  - **Iris**: Round and smooth
  - **IOL**: On the capsular bag
  - **Fundus**: Well-visible and attached

**Assessment**:

- Good postoperative progress in both eyes
- Mild inflammation in the left eye, within the normal range for recovery

**Plan**:
Discharge the patient today.

**Postoperative Medications for Both Eyes**:

- Levofloxacin Hydrate (4x/day)
- Fluorometholone (4x/day)
- Bromfenac Sodium Hydrate(2x/day)

**Case 2: Middle-aged Male with Traumatic Eye Injury**

**Patient Name:** Taro Osaka
**Gender:** Male
**Age:** 45 years
**Date of First Visit:** March 1, 2024

**Diagnosis**

- Penetrating ocular trauma of the left eye

**History of Present Illness**

A 45-year-old male presented to the emergency department with severe pain and significant vision loss in the left eye following a workplace injury. The injury occurred while using a grass trimmer, during which a broken blade fragment penetrated his left eye.

**Past Medical History**

- No significant medical history
- Vaccinated against diphtheria, pertussis, and tetanus (DPT)

**Current Medications**

- None

**Allergies**

- History of anaphylactic shock triggered by shellfish

**Social History**

The patient works as a software engineer in an urban company and lives alone.

**Emergency Department Notes**

**Subjective:**
"I feel pain in my left eye. I’ve never had major illnesses before and have no metal implants in my body."

**Objective:**

- **Vital signs:**
  SpO₂: 98%, HR: 89 bpm, BP: 135/80 mmHg, Temp: 36°C, RR: 16 breaths/min, GCS: 15
- **CT scan findings:**
  Metallic foreign body identified in the globe of the left eye; no evidence of intracranial trauma.

**Assessment:**

- Stable systemic condition
- Suspected ruptured globe

**Plan:**

- Immediate referral to ophthalmology for specialized evaluation and management

**Ophthalmology Evaluation**

**Subjective:**
"I can barely see out of my left eye."

**Objective:**

- **Visual Acuity:**
  - **Right Eye (RV):** 1.2 (1.5x S 0D, C -0.75D, Ax 105)
  - **Left Eye (LV):** Hand motion only
- **Intraocular Pressure (IOP):**
  - RE: 10.6 mmHg
  - LE: 3.6 mmHg
- **Slit-lamp Examination:**
  - **Right Eye :**
    - Conjunctiva: Normal
    - Cornea: Clear
    - Anterior chamber (AC): Deep, no cells
    - Iris: Round and smooth
    - Lens: Normal (NS0)
  - **Left Eye :**
    - Cornea: Vertical laceration across the central region
    - AC: Shallow, hyphema (5 mm)
    - Iris and lens: Partially visible but obscured by corneal laceration; posterior segment was not visualized.
- **CT**: Confirmed metallic foreign body in the orbit.

**Assessment:**

- Penetrating ocular trauma in the left eye
- Corneal perforation with intraocular metallic foreign body

**Plan:**

1. Admit the patient for surgery under general anesthesia on the same day.
2. Start preoperative antibiotics immediately.
3. Administer tetanus toxoid.
4. Perform preoperative blood tests, ECG, and chest X-ray.
5. Surgical procedures to include corneal wound repair, foreign body removal, lens extraction, and removal of vitreous hemorrhage. Address retinal detachment if present.
6. Inform the patient about the possibility of poor visual outcomes despite surgical intervention. In severe cases, enucleation may be required. Explain the potential risk of sympathetic ophthalmia.
7. Discharge will depend on postoperative progress.

**Postoperative Informed Consent (IC):**

The surgery proceeded without complications. A metallic fragment was removed from the globe. Corneal suturing, vitreous hemorrhage removal, and lens extraction were successfully performed. Retinal detachment with an associated retinal tear was treated with laser photocoagulation and silicone oil tamponade. The globe was preserved, though the patient was informed about the risks of infection and sympathetic ophthalmia, which could necessitate enucleation. The patient demonstrated a clear understanding of the explanation.

**Postoperative Progress**

**Hospital Day 2 (March 2, 2024)**
**Postoperative Day 1 (POD1)**

**Subjective:**
"The pain is manageable."

**Objective:**

- **IOP:**
  - RE: 13 mmHg
  - LE: 4 mmHg
- **Visual Acuity (LV):** Hand motion only, accurate light projection in four directions
- **Slit-lamp Findings :**
  - **Right Eye :**
    - normal
  - **Left Eye**
    - Conjunctiva: Marked hyperemia (H++)
    - Cornea: 10-0 nylon sutures in place; no leaks, good wound adaptation
    - AC: Deep, cells (+), fibrin (+)
    - Iris: Mostly round
    - Lens: Aphakia
    - Fundus: Obscured by vitreous hemorrhage

**Assessment:**

- No postoperative complications; no signs of inflammation in the right eye.

**Plan:**

- Postoperative eye drops (LE):
  - Levofloxacin Hydrate　6x/day
  - Cefmenoxime Hydrochloride 6x/day
  - Betamethasone Sodium Phosphate 2x/day
- Intravenous antibiotics (Ceftazidime) 2 g/day

**Hospital Day 3 (March 3, 2024)**
**Left Eye: Postoperative Day 2 (POD2)**

**Subjective:**
"My vision hasn’t improved much yet."

**Objective:**

- **IOP:**
  - LE: 3 mmHg
- **Visual Acuity (LV):** Hand motion only
- **Slit-lamp Findings:**
  - **Right Eye :**
    - normal
  - **Left Eye**
    - Conjunctiva: Hyperemia (H++)
    - Cornea: 10-0 nylon sutures in place; no leaks, good wound adaptation; mild edema and Descemet folds (+)
    - AC: Deep, cells (+), fibrin (+)
    - Iris: Mostly round
    - Lens: Aphakia
    - Fundus: Veiled

**Assessment:**

- Postoperative recovery is satisfactory; no right eye inflammation.

**Plan:**

- Continue postoperative medications as prescribed.
- Plan discharge the next day.
- Postoperative eye drops (LE):
  - Levofloxacin Hydrate　6x/day
  - Cefmenoxime Hydrochloride 6x/day
  - Betamethasone Sodium Phosphate 2x/day
- Intravenous antibiotics (Ceftazidime) 2 g/day

**Hospital Day 4 (March 4, 2024)**
**Left Eye: Postoperative Day 3 (POD3)**

**Subjective:**
"Thank you for taking care of me."

**Objective:**

- **IOP:**
  - LE: 4 mmHg
- **Visual Acuity (LV):** Hand motion only
- **Slit-lamp Findings:**
  - **Right Eye :**
    - normal
  - **Left Eye**
    - Conjunctiva: Hyperemia (H++)
    - Cornea: 10-0 nylon sutures in place; no leaks, good wound adaptation; mild edema and Descemet folds (+)
    - AC: Deep, cells (+), fibrin (+)
    - Iris: Mostly round
    - Lens: Aphakia
- Fundus: Veiled

**Assessment:**

- Postoperative recovery is uneventful; no inflammation in the right eye.

**Plan:**

- Discharge today.
- Continue postoperative eye drops (LE):
  - Levofloxacin Hydrate　6x/day
  - Cefmenoxime Hydrochloride 6x/day
  - Betamethasone Sodium Phosphate 2x/day
- Start oral Levofloxacin Hydrate 500 mg/day.
- Schedule follow-up in the outpatient clinic for further observation.

**Case 3: A 45-Year-Old Woman Presenting with Rhegmatogenous Retinal Detachment**

**Patient Name:** Shiga Yuki
**Gender:** Female
**Age:** 45 years
**First Visit Date:** March 1, 2024

**Diagnosis**

1. Rhegmatogenous retinal detachment (left eye)
2. Bilateral myopic astigmatism

**History of Present Illness**

The patient reported the onset of floaters in her left eye three days prior to consultation. On the day of her visit, she experienced a progressive superior visual field defect in the same eye. She was urgently referred for evaluation by her previous ophthalmologist.

**Past Medical History**

- Atopic dermatitis

**Current Medications**

- Topical steroid ointment (specific details unknown), applied to the face twice daily

**Allergies**

- None

**Social History**

The patient is originally from China and works as a Chinese language teacher. She resides in Japan with her husband and son.

**Objective**

**Visual Acuity (VA):**

- **Right Eye (R):** 1.0 (correctable to 1.5; spherical: -0.50D, cylindrical: -0.50D, axis: 55°)
- **Left Eye (L):** 0.3 (correctable to 1.0; spherical: -0.50D, cylindrical: -0.75D, axis: 105°)

**Intraocular Pressure (IOP):**

- 12 mmHg in both eyes

**Biometric Parameters:**

- **Axial Length:** 24.0 mm (R), 24.2 mm (L)
- **Anterior Chamber Depth:** 3.9 mm (R), 3.78 mm (L)
- **Corneal Endothelial Cell Density:** 2883 cells/mm² (R), 2774 cells/mm² (L)

**Pupillary Reflexes:**

- Direct and consensual reflexes intact bilaterally
- Relative afferent pupillary defect (RAPD): Negative

**Ocular Motility:**

- Normal

**Anterior Segment Findings**

**Right Eye (R):**

- **Conjunctiva:** Conjunctival injection with papillae
- **Cornea:** Clear
- **Anterior Chamber (AC):** Deep, no cells
- **Iris:** Round and smooth
- **Lens:** Nuclear sclerosis grade 1 with anterior capsule opacification
- **Fundus:** Retina attached; peripheral lattice degeneration noted

**Left Eye (L):**

- **Conjunctiva:** Normal
- **Cornea:** Clear
- **AC:** Deep, no cells
- **Iris:** Round and smooth
- **Lens:** Nuclear sclerosis grade 1
- **Fundus:** Inferior bullous retinal detachment with associated lattice tears

**Ancillary Testing:**

- **Fundus Color Photography:** No abnormalities in the posterior pole
- **Optical Coherence Tomography (OCT):** Macular structure intact

**Assessment**

1. Left-eye rhegmatogenous retinal detachment

**Plan**

The patient was admitted for emergency cataract and vitrectomy surgery (L) PEA + IOL + PPV + SF6 gas.

**Postoperative Informed Consent (IC):**
The planned surgery was successfully completed. After cataract extraction and intraocular lens implantation, the retina was reattached. Laser photocoagulation was applied to the retinal tear and drainage site, and the intraocular space was filled with SF6 gas. Postoperative instructions included maintaining a face-down position until the gas dissipated. The patient demonstrated a clear understanding of the explanation.

**Hospital Progress Notes**

**Day 2 (March 2, 2024):**

- **S:** "Things look blurry, but there is no pain."
- **O:** (Left eye)
  - **Intraocular Pressure (IOP):** LT 12 mmHg
  - **Conjunctiva:** Conjunctival hyperemia present; papillary reaction noted.
  - **Cornea:** Bloody keratic precipitates (KPs) observed.
  - **Anterior Chamber (AC):** Deep; presence of inflammatory cells (+).
  - **Lens:** IOL securely fixed.
  - **Fundus (Fds):** Gas fill at 90%; white flecks visible; retina attached.
- **A:** Postoperative course progressing well

4. **P:** Continue face-down positioning; maintain medications (Levofloxacin Hydrate (4x/day), Fluorometholone (4x/day), Bromfenac Sodium Hydrate(2x/day))

**Day 3 (March 3, 2024):**

- **S:** "I feel teary."
- **O:** (Left eye)
  - **Intraocular Pressure (IOP):** LT 11 mmHg
  - **Conjunctiva:** Conjunctival hyperemia present; papillary reaction noted.
  - **Cornea:** Bloody keratic precipitates (KPs) observed.
  - **Anterior Chamber (AC):** Deep; presence of inflammatory cells (+).
  - **Lens:** IOL securely fixed.
  - **Fundus (Fds):** Gas fill at 85%; white flecks visible; retina attached.
- **A:** Postoperative course progressing well
- **P:** Continue current positioning and medications.

**Day 4 (March 4, 2024):**

- **S:** "I'm trying my best to stay face down."
- **O:** (Left eye)
  - **Intraocular Pressure (IOP):** LT 13 mmHg
  - **Conjunctiva:** Conjunctival hyperemia present; papillary reaction noted.
  - **Cornea:** Bloody keratic precipitates (KPs) observed.
  - **Anterior Chamber (AC):** Deep; presence of inflammatory cells (+).
  - **Lens:** IOL securely fixed.
  - **Fds**: Gas fill 75%; retinal redetachment observed at the liquid-gas interface
- **A:** Retinal redetachment
- **P:** Emergency reoperation scheduled.

**Reoperation informed consent:**
The retina was successfully reattached during reoperation, with laser photocoagulation applied. The intraocular space was filled with silicone oil. The patient was informed of the potential for redetachment following the removal of silicone oil. The patient demonstrated a clear understanding of the explanation.

**Day 5 (March 5, 2024):**

- **S:** "I have no eye pain, but my neck is starting to hurt."
- **O:** (Left eye)
  - **Intraocular Pressure (IOP):** LT 10 mmHg
  - **Conjunctiva:** Palpebral conjunctival hyperemia present; papillary reaction noted (+).
  - **Cornea:** Bloody keratic precipitates (KPs) observed.
  - **Anterior Chamber (AC):** Deep; inflammatory cells (+++) present; no oil droplets observed (-).
  - **Lens:** IOL securely fixed.
  - **Fds:** Full silicone oil fill; white flecks visible; retina attached.
- **A:** Post-reoperation course progressing well
- **P:** Continue face-down positioning and current medications. (Levofloxacin Hydrate (4x/day), Fluorometholone (4x/day), Bromfenac Sodium Hydrate(2x/day))

**Day 6 (March 6, 2024):**

- **S:** "Can I go home soon?"
- **O:**
  - **Intraocular Pressure (IOP):** LT 11 mmHg
  - **Conjunctiva:** Palpebral conjunctival hyperemia present; papillary reaction noted (+).
  - **Cornea:** Bloody keratic precipitates (KPs) observed.
  - **Anterior Chamber (AC):** Deep; inflammatory cells (++) present
  - **Lens:** IOL securely fixed.
  - **Fds:** Full silicone oil fill; white flecks visible; retina attached.
- **A:** Postoperative course progressing well
- **P:** Discharge planned for the following day. Continue current medications.

**Day 7 (March 7, 2024):**

- **S:** "Thank you for your care."
- **O:** (Left eye)
  - **Intraocular Pressure (IOP):** LT 9 mmHg
  - **Conjunctiva:** Palpebral conjunctival hyperemia present; papillary reaction noted (+).
  - **Cornea:** Bloody keratic precipitates (KPs) observed.
  - **Anterior Chamber (AC):** Deep; inflammatory cells (+) present
  - **Lens:** IOL securely fixed.
  - **Fds:** Full silicone oil fill; white flecks visible; retina attached.
- **A:** Postoperative course progressing well
- **P:** Discharged with instructions for follow-up care. Outpatient laser photocoagulation for lattice degeneration in the right eye was advised.
- Maintain medications (Levofloxacin Hydrate (4x/day), Fluorometholone (4x/day), Bromfenac Sodium Hydrate(2x/day)
- Postural restrictions are no longer required

**Case 4: A 76-Year-Old Woman Presenting with Corneal Ulcer**

**Patient Name:** Toshiko Mie
**Gender:** Female
**Age:** 76 years
**First Visit Date:** April 1, 2024

**Diagnosis**

1. Infectious corneal ulcer (right eye)
2. Bilateral myopic astigmatism

**History of Present Illness**

A 76-year-old woman presented with pain and redness in her right eye for two days. Symptoms progressively worsened, and she was referred by her previous ophthalmologist.

**Past Medical History**

- Diabetes mellitus
- Hypertension
- Hyperlipidemia
- Chronic heart failure

**Current Medications**

- Pitavastatin Calcium Hydrate1 mg: 1-0-0
- Azosemide 30 mg: 1-0-0
- Alogliptin Benzoate 25 mg: 1-0-0
- Febuxostat 10 mg: 1-0-0
- Empagliflozin 10 mg: 1-0-0
- Furosemide 20 mg: 1-0-0
- Spironolactone 25 mg: 1-0-0
- Edoxaban 30 mg: 0.5-0-0
- Verapamil Hydrochloride 40 mg: 1-0-0

**Allergies:** None

**Social History**

The patient lives alone and is independent in activities of daily living (ADLs).

**O:**

**Visual Acuity (VA):**

- **Right Eye (R):** 0.3 (correctable to 0.5; spherical: -0.50D, cylindrical: -0.50D, axis: 100°)
- **Left Eye (L):** 0.5 (correctable to 1.0; spherical: -0.50D, cylindrical: -0.75D, axis: 110°)

**Intraocular Pressure (IOP):**

- **Right:** 15 mmHg
- **Left:** 16 mmHg

**Anterior Segment Findings**

**Right Eye (R):**

- **Conjunctiva:** Ciliary injection
- **Cornea:** Central infiltrative lesion with epithelial edema and an ulcer evident under fluorescein staining; Descemet's folds (++).
- **Anterior Chamber (AC):** Deep with cells (++); hypopyon present.
- **Iris:** Round and smooth.
- **Lens:** Nuclear sclerosis grade 2.
- **Fundus:** Not visible due to anterior chamber inflammation.

**Anterior Segment OCT (CASIA)**: Corneal thinning observed in the ulcer region; no perforation.

**Left Eye (L):**

- **Conjunctiva:** Normal.
- **Cornea:** Clear.
- **AC:** Deep with no cells.
- **Iris:** Round and smooth.
- **Lens:** Nuclear sclerosis grade 2.
- **Fundus:** Well visible.

**Fundus Photography:**

- Right eye: Imaging not possible.
- Left eye: No abnormalities.

**OCT Findings:**

- Right eye: Imaging not possible.
- Left eye: No abnormalities.

**A:**

Right corneal ulcer.

**P：**

Scraping was performed on the ulcer for specimen collection. Gram, Giemsa, and fungal flora staining were conducted, and cultures were submitted. The patient was admitted for intravenous and topical antibiotic therapy.

**Hospital Course**

**Day 1 (April 1, 2024):**

- **S:** "It hurts."
- **O:**
  - Gram-positive cocci observed in Gram staining.
- **A:** Bacterial keratitis.
- **P:**
  - Topical antibiotics: Cefmenoxime(6x/day), Gatifloxacin Hydrate(6x/day)
  - Eye ointment: Ofloxacin (2x/day)
  - Cycloplegic: Tropicamide (2x/day)
  - IV antibiotics: Ampicillin Sodium/Sulbactam Sodium 1.5 g, twice daily

**Day 2 (April 2, 2024):**

- **S:** "It still hurts."
- **O:**
  - IOP: 12 mmHg (R), 10 mmHg (L)
  - Right eye
    - Conjunctiva: Ciliary injection noted.
    - Cornea: Central infiltration observed, with an ulcer in the same region stained by fluorescein. Surrounding epithelial edema is present, with significant Descemet folds (DF++).
    - Anterior Chamber: Deep, with numerous cells (++). Hypopyon is present.
    - Iris: Round and smooth.
    - Lens: Nuclear sclerosis, grade 2 (NS2).
    - Fundus: Observation is difficult due to anterior chamber inflammation.
- **A:** Stable condition.
- **P:** Continue treatment.

**Day 3 (April 3, 2024):**

- **S:** "The pain has slightly improved."
- **O:**
  - IOP: 10 mmHg (R), 10 mmHg (L)
  - Right eye
    - Conjunctiva: Slight improvement in ciliary injection.
    - Cornea: Marginal reduction in the size of the corneal ulcer.
    - Anterior Chamber: Deep; with cells(++). Hypopyon shows a reduction in severity.
    - Iris: Round and smooth.
    - Lens: Nuclear sclerosis, grade 2 (NS2)
    - Fundus: Slightly visible despite anterior segment findings.
- **A:** Mild improvement.
- **P:** Continue treatment.

**Day 4 (April 4, 2024):**

- **S:** "It feels better."
- **O:**
  - IOP: 12 mmHg (R), 11 mmHg (L)
  - Right eye
    - Conjunctiva: Slight improvement in ciliary injection.
    - Cornea: Reduction in ulcer size.
    - Anterior Chamber: Deep, with inflammatory cells (cell++). Hypopyon ±.
    - Iris: Round and smooth.
    - Lens: NS2.
    - Fundus: Visible.
- **A:** Improvement.
- **P:** Continue treatment.

**Day 5 (April 5, 2024):**

- **S:** "My vision is getting better."
- **O:**
  - IOP: 10 mmHg (R), 11 mmHg (L)
  - Right Eye
    - Conjunctiva: Mild injection.
    - Cornea: Reduction in ulcer size.
    - Anterior Chamber: Deep, with inflammatory cells (cell++). Hypopyon ±.
    - Iris: Round and smooth.
    - Lens: NS2.
    - Fundus: Visible.
- **A:** Improvement trend.
- **P:** Plan discharge timing.

**Day 6 (April 6, 2024):**

- **S:** "Can I go home soon?"
- **O:**
  - IOP: 9 mmHg (R), 10 mmHg (L)
  - Right eye
    - Conjunctiva: Mild injection.
    - Cornea: Near-complete resolution of the ulcer, as confirmed by fluorescein staining.
    - Anterior Chamber: Deep, with inflammatory cells (cell++). Hypopyon ±.
    - Iris: Round and smooth.
    - Lens: NS2.
    - Fundus: Visible.
- **A:** Stable improvement.
- **P:** Discharge scheduled for the next day.

**Day 7 (April 7, 2024):**

- **S:** "Thank you for taking care of me."
- **O:**
  - **VA:**
    - Right: 0.5 (correctable to 0.8; spherical: -1.0D, cylindrical: -0.75D, axis: 120°)
    - Left: 0.5 (correctable to 1.0; spherical: -0.75D, cylindrical: -0.5D, axis: 110°)
  - IOP: 10 mmHg (R), 12 mmHg (L)
  - Corneal ulcer healed; clear conjunctiva.
- **A:** Favorable recovery.
- **P:**
  - Discharged with instructions to continue Cefmenoxime (6x/day)and Gatifloxacin Hydrate(6x/day).
  - Ofloxacin and Tropicamide discontinued.
  - IV antibiotics stopped.
  - Outpatient follow-up for further observation.

**Case 5: A 32-Year-Old Female Diagnosed with Optic Neuritis**

**Patient Information**

- **Name:** Toshiko Nara
- **Gender:** Female
- **Age:** 32 years
- **Date of Initial Visit:** May 11, 2024

**Diagnosis**

- Left optic neuritis

**History of Present Illness**

The patient experienced cold symptoms one month ago. Two weeks prior to the visit, she began noticing progressive vision loss and color perception changes in her left eye, which failed to improve. She was referred to our hospital for further evaluation. She also reported transient worsening of her vision during bathing and physical activity.

**Past Medical History**

- No significant medical history
- No family history

**Current Medications**

- None

**Allergies**

- None

**Social History**

The patient previously worked as a temporary employee at a factory but left her job to care for a family member. She is currently unemployed.

**Ophthalmic Examination**

**Visual Acuity (VA):**

- **Right eye (RV):** 1.5 (not corrected)
- **Left eye (LV):** 0.8 (not corrected)

**Intraocular Pressure (IOP):**

- Right: 15 mmHg
- Left: 20 mmHg
- **Relative Afferent Pupillary Defect (RAPD):** Positive in the left eye
- **Ocular Motility:** Normal
- **Color Vision Test (Ishihara):** 10 errors in the left eye
- **Critical Flicker Fusion (CFF):**
  - Right eye: Disappearance threshold 50.0 Hz, Appearance threshold 50.0 Hz
  - Left eye: Disappearance threshold 23.0 Hz, Appearance threshold 27.0 Hz
- **Goldmann Visual Field Test:** Central scotoma within 10 degrees in the left eye

**Imaging:**

- **MRI with Contrast:** Enhancement observed in the left optic nerve. No findings suggestive of multiple sclerosis (MS) or neuromyelitis optica spectrum disorder (NMOSD) in the brain parenchyma or brainstem.

**Anterior Segment Examination in Both Eyes:**

- **Conjunctiva (Conj):** Normal
- **Cornea:** Clear
- **Anterior Chamber (AC):** Deep, no cells detected
- **Iris:** Round, smooth
- **Lens:** Nuclear sclerosis (NS) grade 0
- **Fundus:**
  - Optic disc in the left eye showed redness and swelling. Retinal vessels and macula appeared normal and attached.

**Laboratory Findings:**

- **Blood Tests:** Normal for CBC, biochemistry, ESR, CRP, ACE, Ca, TPHA
- **Autoimmune Markers:** ANA, RF, anti-SS-A/SS-B, ANCA, anti-MOG antibody, and anti-AQP4 antibody results pending
- **Hepatitis B and C:** No history or markers of infection
- **CSF Analysis:** Normal for oligoclonal bands, myelin basic protein, and IgG index

**A:**

- Left optic neuritis (suspected typical optic neuritis)

**P:**

- Emergency hospitalization initiated for treatment with high-dose corticosteroid pulse therapy.
- Pending laboratory results for anti-MOG antibody and anti-AQP4 antibody to rule out atypical optic neuritis.

**Hospital Course**

**Hospital Day 2 (May 12, 2024):**

- **Subjective (S):** "The vision in my left eye feels about the same as yesterday."
- **Objective (O):**
  - **IOP:** Right 10 mmHg, Left 12 mmHg
  - Left RAPD positive
  - Fundus examination: Persistent redness and swelling of the left optic disc
- **Assessment (A):** Suspected typical optic neuritis
- **Plan (P):** Initiated steroid pulse therapy with Methylprednisolone Sodium Succinate 1 g/day (May 12–14).

**Hospital Day 3 (May 13, 2024):**

- **S:** "My vision seems slightly better."
- **O:**
  - **IOP:** Right 10 mmHg, Left 10 mmHg
  - Left RAPD positive
  - Fundus: Persistent optic disc swelling
- **A:** Left optic neuritis improving on Day 2 of steroid pulse therapy
- **P:** Continued Methylprednisolone Sodium Succinate 1 g/day. Transitioned plan to oral prednisolone (PSL) 30 mg/day starting May 15.

**Hospital Day 4 (May 14, 2024):**

- **S:** "My vision feels about the same as yesterday."
- **O:**
  - **IOP:** Right 9 mmHg, Left 9 mmHg
  - Left RAPD positive
  - Fundus: Persistent optic disc swelling
  - Autoimmune marker results returned negative (ANA, RF, anti-SS-A/SS-B, ANCA).
- **A:** Suspected typical optic neuritis improving; Day 3 of steroid pulse therapy
- **P:** Completed pulse therapy.

**Hospital Day 5 (May 15, 2024):**

- **S:** "I think my vision is better than before treatment."
- **O:**
  - **VA:** Right 1.5 (not corrected), Left 1.0 (not corrected)
  - **CFF:**
    - Right eye: Disappearance 45.0 Hz, Appearance 47.0 Hz
    - Left eye: Disappearance 35.0 Hz, Appearance 37.0 Hz
  - Left RAPD positive
  - Fundus: residual optic disc swelling in the left eye
- **A:** Symptoms improving
- **P:** Planned discharge the following day.

**Hospital Day 6 (May 16, 2024):**

- **S:** "My vision is getting better."
- **O:** Persistent mild optic disc swelling in the left eye, with positive left RAPD.
- **A:** Symptoms improving.
- **P:** Patient discharged with outpatient follow-up. Pending results for anti-MOG and anti-AQP4 antibodies.

**Case 6: A male patient requiring glaucoma surgery**

**Patient Name:** Hyogo Jiro
**Gender:** Male
**Age:** 78 years
**First Visit Date:** 2024.06.01

**Diagnosis:**

- Bilateral Exfoliation Glaucoma
- Bilateral Hyperopic Astigmatism

**History of Present Illness:**
A 78-year-old male referred to our hospital for cataract evaluation. Although not noted in the referral letter, intraocular pressures (IOP) were found to be abnormally high during his initial visit: 34 mmHg in the right eye and 47 mmHg in the left eye.

**Past Medical History:**

- Sleep apnea
- Dyslipidemia
- No significant family history

**Current Medication:** None
**Allergies:** None

**Social Background:**
A retired neurologist.

**S:** ”No pain. I have visual difficulties for several years.”

**O:**
**Visual Acuity:**

- Right eye: 0.15 (corrected to 0.6 with S +2.50D, C -1.25D, Ax 85)
- Left eye: Counting fingers

**Intraocular Pressure:** 34 mmHg (right), 47 mmHg (left)

**Ophthalmic Measurements:**

- Axial length: 22.7/22.4 mm
- Anterior chamber depth: 3.19/3.31 mm
- Corneal endothelial cell density: 2712/2294

**Pupillary light reflex**: Normal

**RAPD**: Negative

**Slit-Lamp Examination:**

- **Both eyes**
  - Conjunctiva: Normal
  - Cornea: clear
  - Anterior Chamber: shallow, pAC/CT=1/2, no cells
  - Gonioscopy (right)open, pigmented, (left) open
  - Iris: Round and smooth.
  - Lens: NS3
  - Fundus: well visible, (Right) Thin neuroretinal rim, (Left) Pale optic disc

**OCT Findings:**

- Right eye: Retinal nerve fiber layer (RNFL) intact.
- Left eye: Diffuse thinning of RNFL.

**Visual Field Testing:**

- Right eye: Intact field with nasal step.
- Left eye: Peripheral field loss, minimal central field remaining.

**A:**

- Untreated Exfoliation Glaucoma and Cataracts in both eyes.
- Left eye is indicated for filtration surgery, but due to age and exfoliation syndrome, trabeculotomy is planned first. Post-surgical outcomes will guide further surgical decisions for the right eye, which has relatively preserved vision. Initiate eye drops in both eyes starting today. The intraocular pressure-lowering effect will be confirmed on the day of admission. Cataract surgery and trabeculotomy will be performed on the left eye.

**P:**

- Initiated eye drops (Carteolol hydrochloride, Latanoprostand, Ripasudil hydrochloride hydrate, and Brimonidine tartrate)in both eyes.
- Follow-up in one week.

**Progress Notes**

**2024.06.08 (Preoperative):**
**S:** "No changes in symptoms."
**O:**IOP: 20 mmHg (right), 14 mmHg (left) with drops.

**A**: Intraocular pressure has been brought within the normal range with eye drops.

**P:** Perform cataract surgery with trabeculotomy on the left eye.

**Postoperative Summary:**

- Successful cataract and glaucoma surgery. Trabecular meshwork incised from 7 to 11 o’clock.

**Hospital Day #2 (2024.06.09):**
**S:** "Left eye feels blurry."
**O:**

- IOP: 18 mmHg (right), 42 mmHg (left).
- **Left eye**
  - Conjunctiva: normal
  - Cornea: Severe epithelial edema
  - Anterior Chamber: Deep, hyphema (4 mm)
  - IOL
  - Fundus: optic disc obscured by blood

**A:** Significant hyphema and elevated intraocular pressure noted.

**P:**

- Performed anterior chamber paracentesis; IOP reduced to 20 mmHg.
- Restarted preoperative drops and initiated oral acetazolamide (500 mg/day, divided doses).
- Continue eye drop treatment for the right eye.

**Hospital Day #2: 2024.06.09, 15:00**

**S:**"Seems like I can see a little better than in the morning, but it’s still blurry."

**O:**

- **IOP**　(under Carteolol hydrochloride, Latanoprostand, Ripasudil hydrochloride hydrate, Brimonidine tartrate, and Acetazolamide):
  - Right eye (R): 13 mmHg
  - Left eye (L): 25 mmHg
- **Examination (L):**
  - Conjunctiva: No pathological findings
  - Cornea: Epithelial edema ++
  - Anterior Chamber (AC): Deep, hyphema 3 mm
  - Lens: IOL well-fixed
  - Fundus (Fds): Not visible due to hemorrhage

**A:**

- Intraocular pressure remains elevated.

**P:**

- Continue eye drops and oral medication.

**Hospital Day #3 (2024.06.10):**
**S:** "Slight improvement in clarity."
**O:**

- IOP(under Carteolol hydrochloride, Latanoprostand, Ripasudil hydrochloride hydrate, Brimonidine tartrate, and Acetazolamide): 16 mmHg (right), 20 mmHg (left).
- Left eye: Mild corneal edema, hyphema (2 mm), disc visible but hazy.

**A:** Decreasing trend in hemorrhage noted

**P:** Continue medications.

**Hospital Day #4 (2024.06.11):**
**S:** "Slightly better, but still blurry."
**O:**

- IOP(under Carteolol hydrochloride, Latanoprostand, Ripasudil hydrochloride hydrate, Brimonidine tartrate, and Acetazolamide): 15 mmHg (right), 18 mmHg (left).
- Left eye: Mild corneal edema, hyphema (2 mm), disc more visible.

**A:** Decreasing trend in hemorrhage noted

**P:** Discontinue oral medication, continue eye drops. Patient likely to be discharged tomorrow.

**Hospital Day #5 (2024.06.12):**
**S:** "Vision is still blurry, but manageable for daily life."
**O:**

- IOP(under Carteolol hydrochloride, Latanoprostand, Ripasudil hydrochloride hydrate, and Brimonidine tartrate): 18 mmHg (right), 17 mmHg (left).
- Left eye: Cornea nearly clear, no hyphema, optic disc fully visible.

**A:** IOP controlled with eye drops alone.
**P:** Continue eye drops and discharge today. Follow-up as an outpatient.
