## Supplement 3 for "Evaluating an LLM-Assisted Workflow for Clinical Documentation: *A Pilot Randomized Controlled Trial on Time and Quality*"

**The Predefined Templates**

Here we describe the Predefined Templates supplied to the LLM assistant, CocktailAI. Although templates would ordinarily be authored by end users, in this study they were prepared in advance by a study team member (T.T.) who was not involved in creating the simulated patient records, in order to reduce participant burden. The templates follow these rules:

- Segments enclosed in {…} are machine-readable directives. The LLM assistant interprets these markers and extracts the specified information directly from the electronic health record (EHR).
- Nesting braces (i.e., {…{…}}) specifies how the extraction should be performed (e.g., required formats, scope, or selection criteria).
- The form {prompt? {…}} passes the inner text to the LLM assistant as a prompt. Use this when you want a flexible, generated response rather than literal data extraction.

| **Document Type** | **Templates** |
| --- | --- |
| Discharge  Summary | Admission Date: {Admission Date {Write in the form of “yyyy Year mm Month dd Day”.}}  Discharge Date: {Discharge Date {Write in the form of “yyyy Year mm Month dd Day”.}}  Discharge Diagnosis:  {Diagnosis {Extract all diagnoses.}}  Allergies:  {Allergy history}  Chief Complaint: {Chief complaint for this admission, or reason for hospitalization}  History of Present Illness: {Present illness {Write in the form of “yyyy Year mm Month dd Day,” and specify Right Eye = R), Left Eye = L), Both Eyes = B).}}  Ophthalmological Medical History: {Ophthalmological medical history}  Systemic Medical History: {Systemic medical history}  Regular Medications: {Eye drops and oral medications at the time of admission.}  Hospital Course: {Prompt? {Briefly describe the course during hospitalization, including surgeries and procedures, using standard style.}}  Discharge Medications: {Discharge medications {List all medications continued from admission to discharge, as well as any new medications added during hospitalization.}}  Condition at Discharge: {Condition of the patient at discharge.}  Discharge Plan: {Discharge plan {Briefly describe the post-discharge plan.}} |
| Discharge Referral | We would like to inform you that the patient referred by your institution has been discharged.  On {Surgery date {Write in the format of “yyyy Year mm Month dd Day.”}}, we performed {Surgical procedure} for {Diagnosis}.  {Postoperative course until discharge {Rewrite the text in a polite style.}}  While we will continue to follow up with the patient at our department, we kindly ask for your continued follow-up during their visits to your institution.  We sincerely appreciate your referral of this valuable case.  We look forward to working with you in the future. |
| Discharge Referral  (No surgery) | We would like to inform you that the patient referred by your institution has been discharged.  The patient was admitted on {Admission date {Write in the format of “yyyy Year mm Month dd Day.”}} and received {Details of treatment during hospitalization} for {Diagnosis} at our hospital.  {Course from admission to discharge {Rewrite the text in a polite style, using “desu/masu” tone.}}  While we will continue to follow up with the patient at our department, we kindly ask for your continued follow-up during their visits to your institution.  We sincerely appreciate your referral of this valuable case.  We look forward to working with you in the future. |
