## Supplement 4 for "Evaluating an LLM-Assisted Workflow for Clinical Documentation: *A Pilot Randomized Controlled Trial on Time and Quality*"

**Evaluation Criteria**

**Domains**

1 **Medical Accuracy**

The comprehensiveness of the information provided in the medical record.

2 **Language**

Text style and vocabulary fit the situation and the intended target group that will read the text.

3 **Conciseness**

The text contains only the strictly necessary information and follows a logical or chronological order.

4 **Presence of Hallucinations**

Any text that is factually incorrect or inconsistent, or unrelated to the medical record.

5 **Validity for clinical use**

The text is deemed as usable in a true clinical scenario dealing with real patient information.

6 **Possibility of harm**

The severity and likelihood of potential harm based on people acting on the text.

**Items**

**【Medical Accuracy Item for Discharge Summary】**

**1-1. Medical Accuracy (Inpatient and Diagnoses)**

Are all relevant Inpatient information and diagnoses correctly provided?

1. There are major factual errors or inaccuracies present (missing or wrong content, misinterpretation of the medical record)
2. The information is generally accurate, but with a few minor errors
3. The information completely accurate, no errors detected
4. Other (please specify)

**1-2. Medical Accuracy (History of present illness)**

Are all relevant aspects adequately addressed?

1. There are major factual errors or inaccuracies present (missing or wrong content, misinterpretation of the medical record)
2. The information is generally accurate, but with a few minor errors
3. The information completely accurate, no errors detected
4. Other (please specify)

**1-3. Medical Accuracy (Hospital Course)**

Are all relevant events and their treatment correctly listed?

1. There are major factual errors or inaccuracies present (missing or wrong content, misinterpretation of the medical record)
2. The information is generally accurate, but with a few minor errors
3. The information completely accurate, no errors detected
4. Other (please specify)

**1-4. Medical Accuracy (Discharge Plan)**

Are all relevant instructions regarding treatment correctly listed?

1. There are major factual errors or inaccuracies present (missing or wrong content, misinterpretation of the medical record)
2. The information is generally accurate, but with a few minor errors
3. The information completely accurate, no errors detected
4. Other (please specify)

**【Medical Accuracy Item for Discharge Referral】**

**1. Medical Accuracy**

　　Are all relevant information correctly provided?

1. There are major factual errors or inaccuracies present (missing or wrong content, misinterpretation of the medical record)
2. The information is generally accurate, but with a few minor errors
3. The information completely accurate, no errors detected
4. Other (please specify)

**【The following are common items】**

**2. Language**

Is the language and style of the text clear enough to be easily understood by its intended audience?

1. The text is not understandable for the desired target group, several corrections are needed.
2. The text is generally understandable for the target group, only minor corrections are needed.
3. The text is understandable for the target group, no corrections are needed.
4. Other (please specify)

**3. Conciseness**

Does the text include only the essential information and present it in a logical or time-sequential manner?

1. Text is poorly summarized (too long or too short), does not follow a chronological or logical order
2. Text is acceptably summarized (could be more extended/concise), generally follows a chronological or logical order
3. Text is accurately summarized
4. Other (please specify)

**4. Presence of Hallucinations**

Are there any factually incorrect or inconsistent or nonsensical text (known as hallucinations) ?

1. Yes
2. No

**5. Approval for clinical use**

Is the text deemed as usable in a true clinical scenario dealing with real patient information?

1. Not approved for clinical use
2. Needs correction, but could be used for clinical use
3. Approved for clinical use
4. Other (please specify)

**6-1. Possibility of harm (extent)**

What is the extent of possible harm?

1. Death or severe harm
2. Moderate or mild harm
3. No harm
4. Other (please specify)

**6-2. Possibility of harm (likelihood)**

What is the likelihood of possible harm?

1. High
2. Medium
3. Low
4. Other (please specify)

**7. How would you rate the overall quality of the document, 0 being the lowest quality**

**and 10 being the highest possible quality?**

　___________

**8. Do you think this document belongs to the Clinician-in-the-loop group, the Clinician-only**

**group or the LLM-only group?**

1. Clinician-in-the-loop group
2. Clinician-only group
3. LLM-only group
